# The Dynamics of SARS-CoV-2 Infectivity with Changes in Aerosol Microenvironment

**DOI:** 10.1101/2022.01.08.22268944

**Authors:** Henry P. Oswin, Allen E. Haddrell, Mara Otero-Fernandez, Jamie F.S. Mann, Tristan A. Cogan, Tom Hilditch, Jianghan Tian, Dan Hardy, Darryl J. Hill, Adam Finn, Andrew D. Davidson, Jonathan P. Reid

## Abstract

Understanding the factors that influence the airborne survival of viruses such as SARS-CoV-2 in aerosols is important for identifying routes of transmission and the value of various mitigation strategies for preventing transmission. We present measurements of the stability of SARS-CoV-2 in aerosol droplets (∼5-10µm equilibrated radius) over timescales spanning from 5 seconds to 20 minutes using a novel instrument to probe survival in a small population of droplets (typically 5-10) containing ∼1 virus/droplet. Measurements of airborne infectivity change are coupled with a detailed physicochemical analysis of the airborne droplets containing the virus. A decrease in infectivity to ∼10 % of the starting value was observable for SARS-CoV-2 over 20 minutes, with a large proportion of the loss occurring within the first 5 minutes after aerosolisation. The initial rate of infectivity loss was found to correlate with physical transformation of the equilibrating droplet; salts within the droplets crystallise at RHs below 50% leading to a near instant loss of infectivity in 50–60% of the virus. However, at 90% RH the droplet remains homogenous and aqueous, and the viral stability is sustained for the first 2 minutes, beyond which it decays to only 10% remaining infectious after 10 minutes. The loss of infectivity at high RH is consistent with an elevation in the pH of the droplets, caused by volatilisation of CO_2_ from bicarbonate buffer within the droplet. Three different variants of SARS-CoV-2 were compared and found to have a similar degree of airborne stability at both high and low RH.

**Significance:** The aerosol microenvironment is highly dynamic exposing pathogens, such as the SARS-CoV-2 virus when exhaled in respiratory aerosol, to extreme conditions of solute concentration, pH and evaporative cooling. Yet surviving this environment is a key step in the transmission of such pathogens. Understanding the impact that airborne transport has on pathogens and the influence of environmental conditions on pathogen survival can inform the implementation of strategies to mitigate the spread of diseases such as COVID-19. We report changes in the infectivity of the airborne virus over timescales spanning from 5 s to 20 minutes and demonstrate the role of two microphysical processes in this infectivity loss: particle crystallisation and aerosol droplet pH change.

## Introduction

The ongoing coronavirus disease 2019 (COVID-19) pandemic has demonstrated the requirement for an improved understanding of the factors that govern the relative importance of different modes of transmission of respiratory pathogens, including the parameters that influence droplet, fomite, and airborne transmission. Indeed, shortcomings in our understanding have prolonged the debate surrounding the likelihood of airborne transmission of severe acute respiratory syndrome coronavirus 2 (SARS-CoV-2) (1–3), with consequences for the implementation of non-pharmaceutical interventions and mitigation strategies such as physical distancing, the wearing of face coverings and the use of ultraviolet germicidal irradiation. Currently, epidemiological evidence (4–6), air sampling studies (7), and animal-model studies (8) are broadly consistent with transmission dominated by close contact with infected individuals. While transmission over longer distances has been documented, it tends to be rare and in preventable circumstances (9), such as prolonged exposure in poorly ventilated rooms (10, 11).

Reports of the airborne stability of SARS-CoV-2 consistently indicate that the half-life associated with the decay in viral infectivity is on the order of hours in surrogates of respiratory aerosols (12–15). However, a detailed understanding of the processes that govern the airborne longevity of viruses, and how infectivity is affected by basic environmental conditions such as relative humidity (RH) and temperature, is required. More specifically, there is little clarity on the impact of environmental conditions on the microenvironment within an airborne droplet, and the interplay between this microenvironment and the stability of pathogens. Improved models of the physicochemical properties of respiratory aerosol and the processes that transform particle size, moisture content, composition and phase are essential to provide clearer insights into the relative risks of airborne transmission in different environments and the potential benefits of mitigation measures to reduce transmission. Indeed, it should be recognised that transformation processes lead to transient changes in properties (e.g. surface enrichment in salts during evaporation following droplet exhalation) that can have distinct impacts on infectivity from the steady state equilibrium properties that persist over longer time periods during airborne transport (e.g. an equilibrated salt concentration).

The microenvironment within an airborne droplet is multifarious and notoriously difficult to study (16), and is further complicated by the presence of organic macromolecules and microorganisms (17). While the vast majority of indoor aerosol originate from sources such as candles, dust, outdoor air pollution and food cookers (18), respiratory pathogens are transmitted in exhaled aerosol that can span from 100 nm to 100 μm diameter and have emission rates as low as 10 particles s^-1^ when breathing (19, 20). Regardless of the expiratory activity that generates respiratory aerosol (e.g. coughing, speaking (20, 21)), the high surface area-to-volume ratio of the emitted particles facilitates rapid equilibration to the surrounding gas phase composition (Figure S1A) (22). In particular, the equilibration of the water activity within the droplet to the surrounding RH impacts the physicochemical conditions experienced by microorganisms present within the aerosol. Aqueous respiratory droplets at the point of exhalation start with a very high-water activity (approximately 0.995) (23) consistent with equilibration with the high RH within the respiratory tract, but must adjust to equilibrate with the indoor humidity, which is typically within the range 20 – 60% (24–26). Under most conditions, exhaled aerosol droplets rapidly lose both moisture and heat through evaporation, with large concomitant changes in volume and temperature as they establish an equilibrium with the indoor environment.

Not only does the loss in water lead to an increase in solute concentrations during evaporation, but the absence of heterogenous nucleation sites (i.e. a surface) leads to supersaturated solute concentrations than cannot be achieved in the bulk solution phase or in sessile droplets deposited on surfaces. At sufficiently low RH (e.g. below 45% for saline solution droplets), the supersaturation of solutes can be sufficient to induce homogenous nucleation (27–29), leading to efflorescence (crystallisation) of the droplet and the formation of a dry particle. Furthermore, during the initial period of droplet evaporation, the rates of diffusion of microorganisms within the droplet can be significantly slower than the rate at which the droplet surface recedes, leading to their exclusion to the near-surface region of the droplet. Given that the physicochemical conditions at the surface of the droplet can be different to the core (e.g. surface enrichment in solute concentration), establishing the distribution of microorganisms within a particle may be crucial to understanding the impact of aerosol microphysics on their longevity.

Once the moisture content of the aerosol has decreased to establish equilibrium with the ambient environment, the decay in microorganism survival may be regulated by steady state microphysical properties. In particular, the typical range in ambient RH is consistent with equilibrated solute concentrations that are supersaturated in the exhaled aerosol. Although the mechanism remains unclear, high salt concentrations may inactivate viruses by damaging the viral RNA (30, 31). With high contents of organic macromolecules, phase-separated particles with organic and inorganic rich domains or amorphous particles containing trapped moisture may form, potentially enhancing viral and bacterial survival. Further, the pH of aerosol particles is RH, size and composition dependent, and the pH of aerosol droplet surfaces may be different from the droplet bulk (32). Indeed, predicting the evolving aerosol pH is challenging, particularly when the facile partitioning of water soluble acidic and basic components from the ambient environment is considered, even before the influence of aerosol pH on microorganism survival is considered (33).

One of two laboratory strategies can be adopted to assess the airborne stability of a pathogen and the interplay of stability with aerosol microphysics. The approach must either be capable of simulating every aspect of the real-world environment in which transmission occurs, or sufficient control over the conditions must be achieved such that the influence of individual processes and properties on survival can be assessed independently. Goldberg rotating drums (34) have been widely used over many decades to assess airborne pathogen stability and have been used to investigate the airborne survival of SARS-CoV-2. More specifically, studies have examined the dependence of infectivity on time (20 minutes – 16 hours), RH (40% - 70%) and the presence of UVC light (12–15, 35), with measurements in aerosols composed of cell culture media (DMEM and MEM) and artificial saliva. All studies concentrate on equilibrated particle sizes of ∼5 μm (mass median aerodynamic diameter). A nebuliser is used to generate a cloud of aerosolised pathogen that is suspended by the rotation of the drum. The initial environmental conditions within the drum can be controlled by mixing the output of the nebuliser with a flow of humidity and temperature-controlled air. However, operation with stable environmental conditions can be challenging; for example, as the droplets evaporate and equilibrate to the set humidity, the water they release can cause the humidity within the drum to increase (see for example the report of Smither et al. (13)). In addition, dynamic changes in liquid water content within the freshly nebulised aerosol cloud do not replicate the very rapid changes that can accompany the extremely low concentrations of exhaled aerosol. This precludes any study of short-term decreases in pathogen viability that may be critical to understanding close contact transmission and the immediate consequences of exhalation on microbe survival.

We have previously reported a unique approach to the study of infectious aerosol and the interplay between aerosol microphysics and pathogen survival, using complementary aerosol analysis techniques to assess the underlying mechanisms that govern the airborne longevity of pathogens (36, 37). The aerosol stability of viruses and bacteria is investigated using the CELEBS technique (Controlled Electrodynamic Levitation and Extraction of Bioaerosols onto a Substrate) (36–38). In CELEBS (Fig. S1B), a small population (<20) of near identical monodisperse droplets containing bacteria or viruses are trapped within an electric field, while a constant flow of air prevents the accumulation of released water around the droplets. Loading droplets into the CELEBS takes < 0.1 s and there is no physical loss of droplets over time. Thus, assessment of viability of suspended microbes within droplets can be made after periods of suspension varying between less than 5 seconds to many hours. These longevity measurements can then be contextualised with detailed measurements of the dynamic changes in the physicochemical properties of droplets generated the exact same way in an instrument referred to as the Comparative Kinetic-Electrodynamic Balance (CK-EDB) (36, 39–43). The CK-EDB uses the same piezoelectric droplet-on-demand dispensers as the CELEBS to generate droplets, with particles captured in the path of a laser within a flow of humidity and temperature-controlled air (Fig. S1C). The elastic light scattering pattern can be used to infer the size and structure of these droplets within the same environmental conditions as those used in CELEBS.

By coupling measurements of the physicochemical properties of the droplets (CK-EDB) with the downstream biological effects (CELEBS), the systematic exploration of hypotheses regarding the inactivation mechanisms of viruses and bacteria is possible. In this study, we apply this approach to the study of SARS-CoV-2 survival in airborne droplets of cell culture medium, examining the survival over timescales spanning from <20 s, commensurate with the evaporation of freshly exhaled aerosol, through to 20 minutes. By studying the physicochemical changes that take place in the droplet and exploring how these changes impact the infectivity of the virus, we elucidate the effect of the airborne environment on SARS-CoV-2. This study provides novel insights into the potential influence of environmental conditions on COVID-19 transmission.

## Results

### The airborne infectivity of SARS-CoV-2 declines over the first 20 minutes following aerosolisation

The infectivity of SARS-CoV-2 contained in MEM 2% FBS was measured over the course of 20 minutes of levitation in CELEBS at both low (40%) and high (90%) RH (Fig. 1A). A decrease in infectivity at low RH occurs almost immediately, falling to an average of 54% within 5 seconds of generation. Interestingly, although the initial loss in infectivity at low RH is almost instant, the virus infectivity then remains more stable, only decreasing an average of 19% over the next 5 minutes. At high RH the reduction in infectivity following aerosolization is more gradual with a steady loss of infectivity of 48% within the first 5 minutes. The decay in survival appears to plateau at both RHs after 10 minutes and the difference between infectivity in aerosol particles suspended at the two RHs diminishes over time, until survival at the two RHs is indistinguishable after 20 minutes. Further research will be required to explore for how long the apparent plateau continues, but it is possible that this slowing down of the viral decay is responsible for the longer half-lives reported in previous Goldberg rotating drum studies (12, 14). It is unlikely that the rapid initial decay in virus infectivity would be observable in a rotating drum due to the relatively long times required to load the drum. The SARS-CoV-2 airborne decay rates reported by previous studies most likely come from this later, flatter portion of the curve.

**Figure 1.**
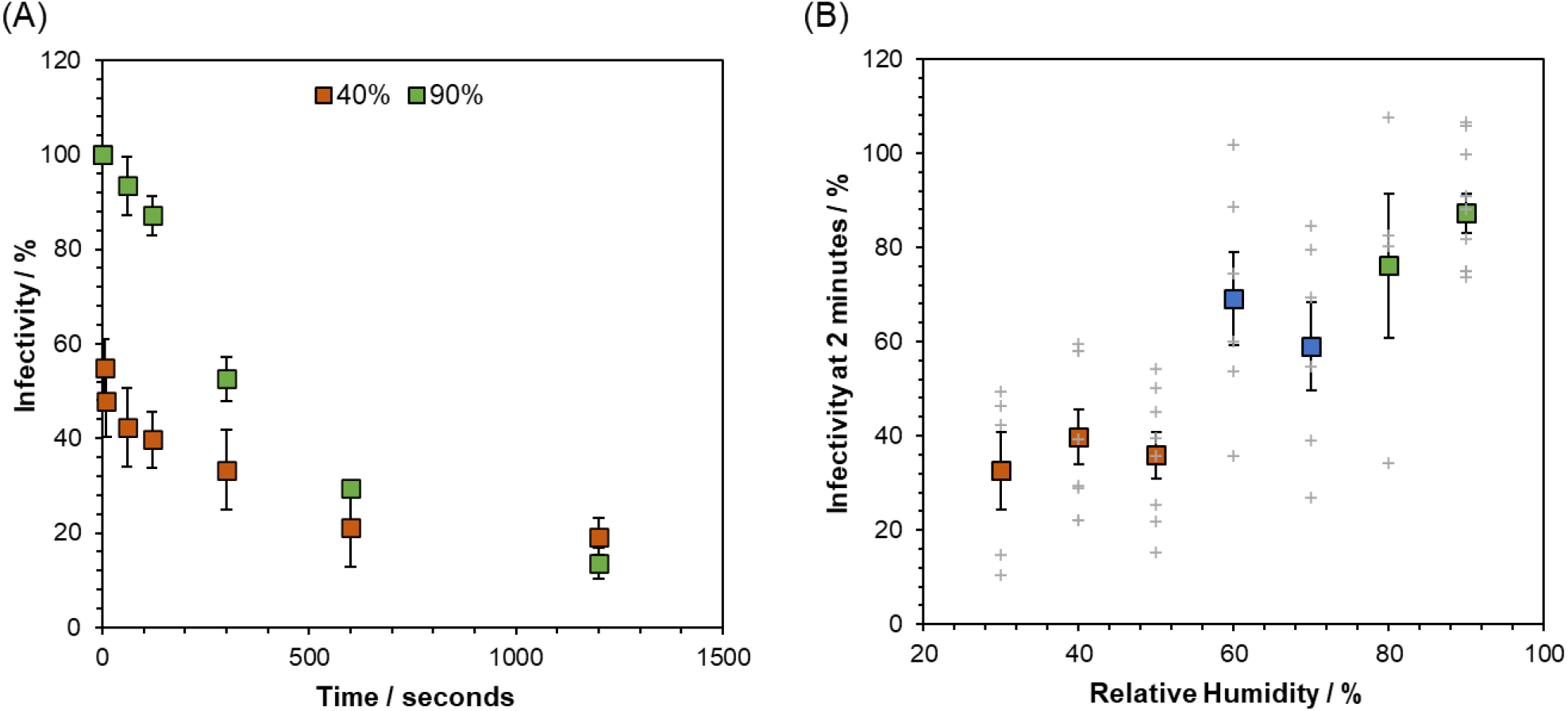
The Short-Term Airborne Decay of SARS-CoV-2. Datapoints are the mean of several measurements (typically >4) and error bars show the standard error. Measurements were carried out at room temperature 18 – 21°C. (A) The percentage infectivity of SARS-CoV-2 REMRQ0001 as a function of time levitated in CELEBS at 40% RH (orange) and 90% RH (green). (B) Curve showing the impact of RH on the percentage infectivity of SARS-CoV-2 REMRQ0001 after 2 minutes of levitation in CELEBS. The larger coloured square points show the mean, with the error bars showing the standard error. Grey crosses show the results of individual measurements.

To more fully characterise the dependence of the infectivity of SARS-CoV-2 on RH, the RH was varied from 30-90 % and the remaining infectivity at 2 minutes measured. Previous studies have reported little dependence of the infectivity decay rate on RH within the uncertainty of the measurements (13, 15). However, we observe a clear relationship between the short-term viability of SARS-CoV-2 and RH (Fig. 1B). Between 30 and 50% RH, the infectivity typically declines within this short time frame to between 30% and 40% after 2 minutes of levitation. At RHs of 80% and above, the virus is far more stable, with infectivity rarely falling below 80% after 2 minutes. The residual infectivity between 60% and 70% RH is highly variable, sometimes falling to similar levels to those observed at the lower RHs and sometimes showing almost no decrease; we shall return to this variability in a later section.

The rapid decay in infectivity reported here, with an observed half-life of on the order of seconds to minutes, has not been reported previously. However, consistent with the majority of previous studies, these survival decays have been measured in virus culture directly and it should be remembered that the aerosol composition (MEM 2% FBS) is different from real exhaled respiratory fluids, including saliva, sputum and other respiratory fluids. Thus, we now investigate the causative mechanisms driving the decay of SARS-CoV-2 in airborne MEM 2% FBS in order to better understand the relevance of these measurements to the transmission of SARS-CoV-2.

### Airborne droplets of MEM show complex phase behaviour during evaporation

To provide insight into the underlying mechanisms that drive the observed airborne loss of SARS-CoV-2 infectivity, the microphysical changes (depicted in Figure S1A) taking place in the droplets hosting the virus were explored in real time and *in situ* using the CK-EDB with a time-resolution of <100 ms (36, 39–43). For context, the phase changes that occur during the evaporation of aqueous sodium chloride at an RH below the efflorescence threshold are shown in Figure S2. When efflorescence occurs at low RH, the crystallisation of the salt is exothermic, resulting in an increase in the droplet temperature and a concomitant transient increase in the evaporation rate. This increase in evaporation rate characteristic of efflorescence is best observed in changes in the intensity of the total light scattered by the particle. By comparison, Mie scattering calculations from the angularly resolved light scattering pattern can be used for precise estimation of the droplet size and can provide other insights into the physical transformations of the particle, such as the formation of numerous submicron crystals dispersed within the host liquid droplet and the point at which the particle ceases to be spherical (44).

For the viral longevity measurements in this study, the virus was suspended in MEM 2% FBS, the tissue culture medium used in the initial growth of the virus on Vero cells. The relatively low viral titres obtained with SARS-CoV-2 (12, 45) culture prevented dilution into other solutions, constraining longevity experiments to the starting stock solution. We avoided concentrating the virus stocks using methods such as ultracentrifugation and tangential flow filtration to avoid any impact these processes might have on the stability of the virus, which could then introduce ambiguity into the interpretation of the longevity data. MEM is a complex solution containing a range of inorganic salts and organic components such as proteins, amino acids, and various sugars. The composition is made more complicated and uncertain through the addition of an animal extract (foetal bovine serum - FBS). Saliva is also a complex mixture of inorganic and organic components, with many solutes at similar concentrations to those found in MEM. Saliva has average NaCl and NaHCO_3_ concentrations of 6.4 and 3.7 g/L respectively (46–48) while MEM typically has concentrations of 6.8 and 2.2 g/L. It should be noted though that the composition of saliva can vary significantly from individual to individual, and over the course of a respiratory infection (46, 47, 49–51).

To better understand the response of aerosols, formed from the complex mixture of components typical of cell culture media and respiratory secretions, to the airborne environment, the drying kinetics of droplets containing MEM 2% FBS were studied using the CK-EDB. Evaporation curves for droplets of MEM 2% FBS levitated at a range of RHs are shown in Figure 2A. At an RH of 51% and below, changes in the overall light scatter intensity typical of efflorescence were observed (Fig. S3), with the droplets crystallising in less than 5 seconds from generation. At a measurement RH of 67%, efflorescence was not observed although the recorded Mie scattering profile indicates that the particles are no longer spherical, potentially forming inhomogeneous amorphous semi-solid particles (Fig. 2a). Indeed, at 78% RH, variability in the outcome of the dynamics and phase transformation of the aerosol was observed: particles initially underwent a phase change (possibly with the formation of inclusions) that was sometimes reversible, reforming a homogenous spherical particle at later time. At RHs of 85% and above, particles mostly remained homogenous aqueous spheres. The dependence of the apparent final particle structure on RH is summarised in Figure 2B. At the extremes of RH, particles of consistent phase were formed following drying and equilibration, with crystalline or spherical homogenous solution droplets resulting at low and high RH, respectively. At intermediate RHs, variability in the physical state of the equilibrated particle was observed, mirroring the greater variability in the remaining infectivity of SARS-CoV-2 at 2 minutes across these RHs (Fig. 1B). We shall return to a fuller explanation of the phase behaviour of the droplets at these intermediate RHs in a later section. These results suggest that the phase of the particle following the equilibration into the gas phase RH directly impacts the short-term infectivity of the virus.

**Figure 2.**
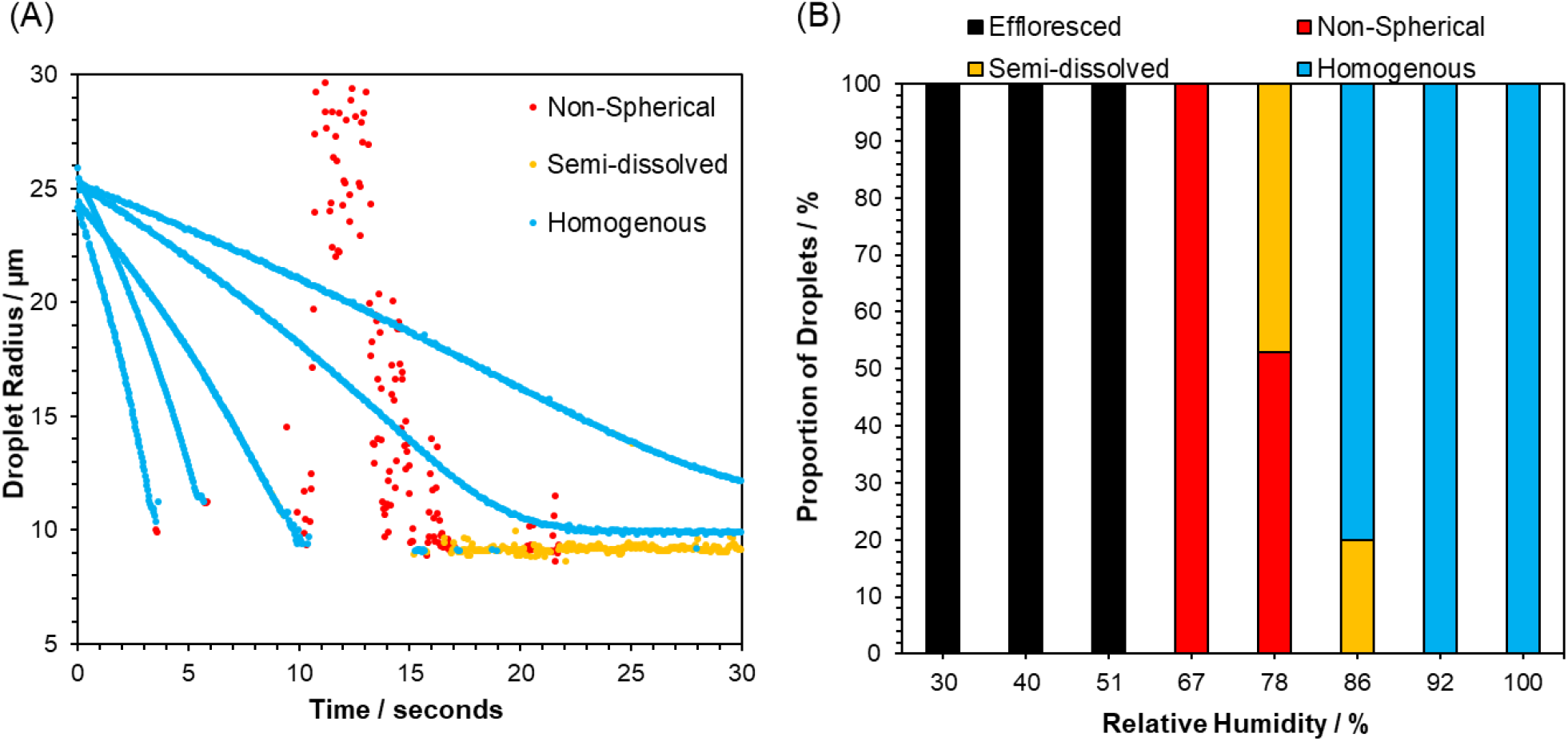
The Microphysics of Airborne MEM Droplets. (A) Mie scatter evaporation profiles of MEM 2% FBS generated by a droplet dispenser and levitated in the CK-EDB at different RHs (51%, 66.8%, 78.2%, 86%, 92%, left to right). Blue indicates a homogenous spherical droplet, yellow indicates the presence of inclusions within the droplet, red indicates a non-spherical particle (note that size estimates become inaccurate for non-spherical particles). (B) Proportion of particle morphologies formed by MEM 2% FBS at different RHs. Multiple levitations were carried out at each RH and there was variation in the final morphology of the particles formed. The frequency of the formation of each particle type is shown for the RHs studied, with black indicating efflorescence, red indicating a non-spherical particle, yellow indicating a semi dissolved particle, and blue indicating an aqueous homogenous particle.

The relationships between the RH, the rate of evaporation and the volume change during drying for aqueous MEM droplets are shown in Figure S4. The solute molarities increase from their initial values by around 10-fold when droplets evaporate into a gas phase at 92% RH and 25-fold at 78.2%, as reflected by the change in droplet radius and, thus, volume. Below this RH, inclusion formation (likely by some of the solute components crystallising from solution) precludes an accurate estimation of the degree of supersaturation achieved within the remaining liquid phase. Although equilibration timescales are size dependent (smaller droplets would be expected to reach equilibrium much faster), the overall increase in solute concentration is size independent.

During equilibration to the ambient RH, the surface of an evaporating droplet can become enriched with larger solutes and suspended matter if the rate at which the surface is receding (κ, m^2^ s^-1^) is faster than the rate of diffusional mixing (reflected in the diffusion constant, *D_i_*, m^2^ s^-1^) (52, 53). This competition is characterised by the Peclet number, *Pe_i_*, for component *i*:

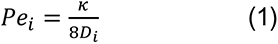

By comparing the evaporation rates reported in Figure 2C with previously reported diffusion coefficient for a typical virus in water (54, 55), the Peclet number for SARS-CoV-2 in MEM 2% FBS can be estimated. In all cases and for all temperatures studied here, the initial Peclet number for SARS-CoV-2 at the starting droplet water activity can be assumed to be in the range 0.5 - 5, showing marginal surface enrichment at most (56). As the water content diminishes during evaporation, particularly when drying into low RH, the increasing solute concentrations may slow the diffusion of the virus and may lead to surface segregation, although we do not account for this here. Indeed, Peclet numbers for more highly diffusing solutes will be <<1 and can be assumed to show only marginal surface enrichment at the lowest RHs and highest temperatures; for example, at a Peclet number of 0.2, drying aqueous sodium chloride droplets show a transient enrichment in surface salt concentration of ∼20% above the droplet core concentration for similar sized droplets (57). As a consequence, the distribution of viruses in evaporating aerosol droplets can be anticipated to remain fairly uniform, at least when compared to the much slower transport of much larger bacteria (36).

### Efflorescence enhances loss of infectivity in aerosol at low RH

The loss of infectivity at low RHs appears to be consistent with observations of a change in phase state for the airborne droplet with a reproducible decrease in infectivity observed when efflorescence occurs. However, it remains unclear whether the efflorescence event itself impacts the infectivity of the virus. To confirm the correlation with phase behaviour further, the RH was cycled above and below the efflorescence/deliquescence points twice during the levitation of droplet populations, with the infectivity measured before and after each efflorescence event (Fig. 3A). Initially levitation at a RH above efflorescence (∼75 %) for 60 seconds resulted in a reduction in infectivity to ∼50 %. On a separate population of droplets, the RH was transiently reduced from 75 % to 40 % for a period of 10 s, sufficient time to drive efflorescence, before being raised to 75 % and the droplets collected.

**Figure 3.**
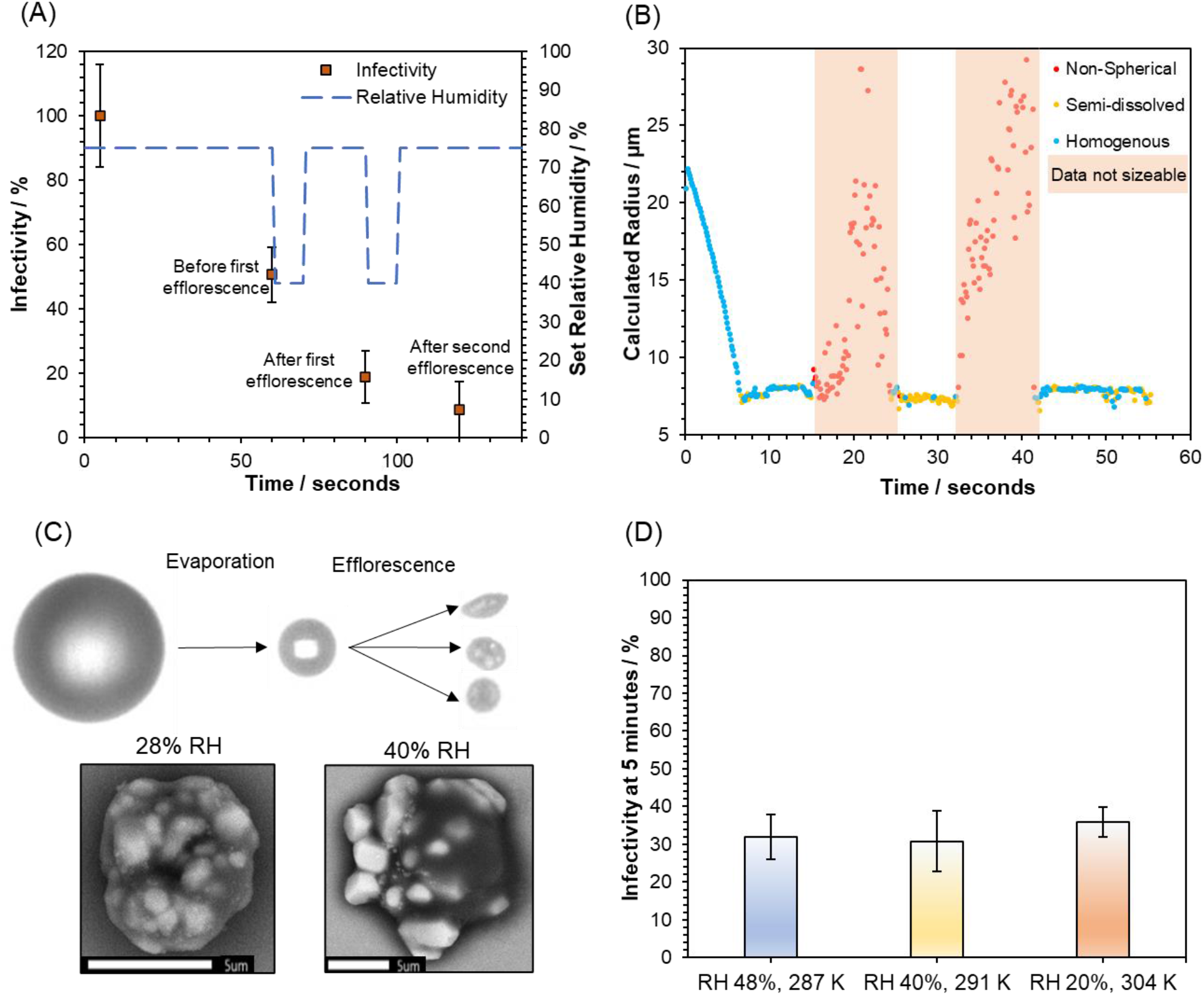
The Role of Efflorescence in SARS-CoV-2 Airborne Loss of Infectivity. (A) Infectivity curve for SARS-CoV-2 (REMRQ0001) levitated at fluctuating RH. The blue dotted line plotted against the left-hand y-axis shows the changes in the set RH throughout the experiment, whilst the orange datapoints show the % infectivity (mean of 4 measurements with error bars showing the standard error). Annotations have been added to datapoints for clarity. (B) CK-EDB measurements showing the phase behaviour of levitated MEM droplets as the RH is cycled between 75% and 40%, in a similar manner to the survival measurements shown in 4A. RH is initially set at 75%, lowered to 40% at 14 seconds, raised to 75% at 24 seconds, lowered to 40% at 32 seconds, and finally raised to 75% at 41 seconds. Structural information about the droplet is denoted by the colour of the data points. Blue when the droplet is homogenous, yellow when solid inclusions are present, red when the particle is no longer spherical (note that size data becomes inaccurate for non-spherical particles and reported sizes are only included to illustrate this). (C) Images showing the changes in particle morphology that take place whilst MEM 2% FBS is airborne. The top images are from the FDC and showing the initial droplet generated by the dispenser on the left, the droplet after 1.6 seconds of evaporation at 28% RH in the centre, and three different particles after they have undergone phase change on the right. The bottom shows two SEM images of droplets collected after airborne efflorescence has taken place at 28% RH (left) and 40% RH (right). The scale bar is 5 µm long. (D) % Infectivity of SARS-CoV-2 (REMRQ0001) measured after levitation for 5 minutes at three different temperatures and RHs. Bars show the mean of 5 measurements with error bars showing the standard error.

The infectivity at this point had fallen to ∼19%. A second efflorescence cycle, reducing the RH to 40 % again, and then raising it before sampling the droplets, resulted in an average infectivity of ∼9%.

In summary, two efflorescence events resulted in a >90% decrease in percentage infectivity in just 2 minutes, lower than in any measurement in which the RH was maintained at a constant value. In addition, the infectivity dropped to below 50% of the preceding infectivity measurement at each efflorescence event (37% for the first efflorescence, 46% for the second), corresponding to the rapid initial decrease seen at 40% RH (Fig. 1A). A CK-EDB measurement of a levitated MEM droplet, in which the RH was cycled between the same values as in the CELEBS survival measurement, is shown in Figure 3B. This data confirms that when a cycle of evaporation and efflorescence, redissolution, efflorescence and redissolution occurs as the RH is cycled between 75, 45, 75, 45 and 75 %. As in previous CK-EDB measurements of MEM, the particles were predominantly aqueous the higher RH but with some inclusion formation. For all measurements in Figure 3A, the droplets were deposited at high RH, meaning they were in a dissolved solution phase. This indicates that the efflorescence driven loss in infectivity did not arise from a physical sequestration of the virus in non- redissolving salt crystals, but reflected an infectivity impairing alteration to the virus itself.

The consistency in the infectivity reduction induced by efflorescence, even when multiple efflorescence events take place in the same droplet population, demonstrates that there is no inherent property of individual virions that protects them from the crystallisation event. The factor that determines whether an individual virion retains infectivity post efflorescence must instead depend on the local conditions in the vicinity of each individual virion. It was possible to image the evaporation and efflorescence of airborne MEM 2% FBS at 40% RH using a falling droplet column (Fig. 3C). In flight, there is considerable variability in the morphology of the MEM particle immediately after crystallisation. The dried MEM 2% FBS droplets following efflorescence in the falling droplet column can be collected and imaged in more detail using scanning electron microscopy (SEM) (also Fig. 3C). These images of the effloresced media reveal that some of the particle is crystalline whilst some is not. Thus, it is possible that whether or not the virus is in the crystallised fraction of a particle determines its stability following efflorescence. Interestingly, the salt crystals formed became smaller and more numerous as the RH is lowered (Fig. S5), consistent with previous work that has shown that there is a greater propensity for nucleation when droplets are dried at higher rates leading to more nucleation events and smaller final crystals forming a larger composite particle (58).

Changing the temperature of the air around the droplets but maintaining the RH below the efflorescence point does not significantly impact the observed loss of infectivity (Fig. 3D). This provides further evidence that the mechanism driving the loss of infectivity is a physical process such as efflorescence rather than a thermodynamically driven chemical process, such as the rate at which the solute concentrations increase during the evaporation process. The temperature change marginally alters the timepoint at which efflorescence occurs, but the droplets all effloresce within 25 seconds for all three temperatures reported here, well before the 5-minute point at which droplets were sampled and infectivity measured.

### Airborne longevity appears similar for different SARS-CoV-2 variants

Most measurements in this study were carried out using SARS-CoV-2 isolated early in the pandemic (SARS-CoV-2/human/Liverpool/REMRQ0001/2020 (REMRQ0001)). We compared the data from this variant to CELEBS measurements with two others to determine if changes in the structure of SARS-CoV-2 could have an impact on its response to the airborne environment. At 5 minutes, a decrease in infectivity was observed at both at 40% and 90% RH for REMRQ0001, providing the optimum time to resolve any differences in aerostability. At both 40 % and 90 % RH, no significant difference was observed between REMRQ0001, B.1.1.7 (the Alpha variant), and a mutant of the SARS-CoV-2 isolate England/2/2020 which has the same Spike protein sequence as REMRQ0001 except that the furin cleavage site is deleted (designated BriSΔ) (59, 60), Figure 4. It is possible that if this comparison is expanded to cover a broader range of times and conditions, differences between these variants will be observable. However, based on these measurements it does not appear that the deletion in BriSΔ, or the array of mutations throughout B.1.1.7, result in readily observable changes in the airborne longevity of the virus when compared to REMRQ0001. There is no reason to believe that the measurements in this study using REMRQ0001 are not representative of later circulating variants of the virus.

**Figure 4.**
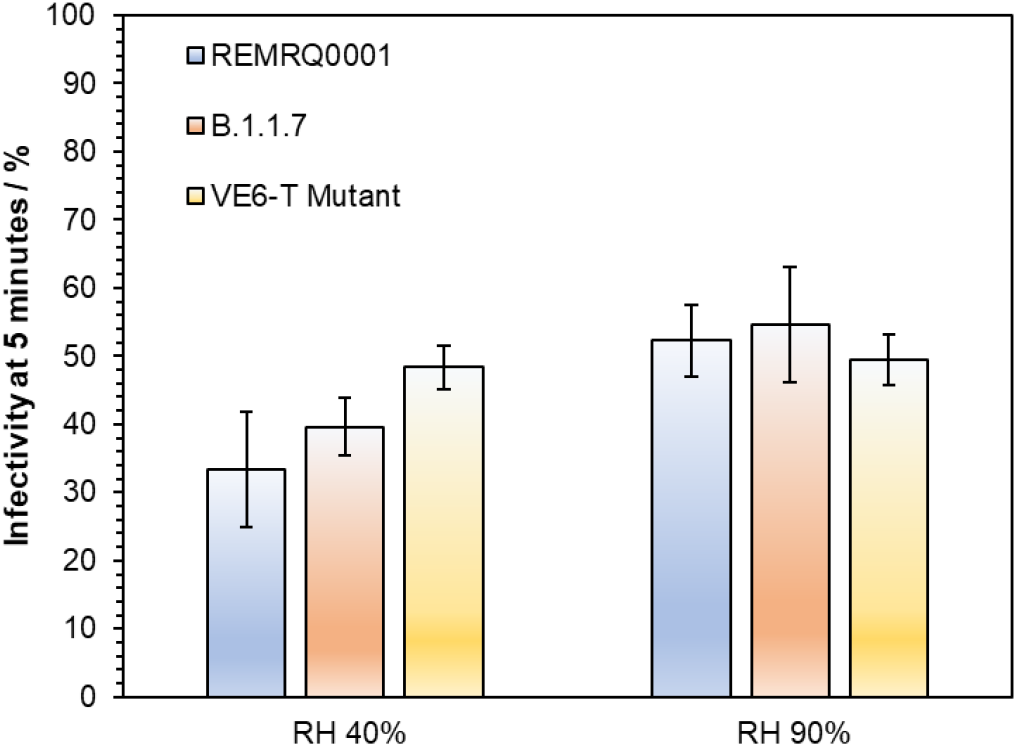
The Influence of SARS-CoV-2 Strain on Airborne Stability. Infectivity of three different variants of SARS-CoV-2 (blue bars for REMRQ0001, orange bars for B.1.1.7, and yellow bars for BrisSΔ). Infectivity is compared after 5 minutes of levitation at 40% and 90% RH, 18°C. At 40% RH, N=5 for REMRQ0001, N=8 for B.1.1.7, and N=4 for BrisSΔ. At 90% RH, N=7 for REMRQ0001, N=11 for B.1.1.7, and N=7 for BrisSΔ. Bars show the mean, error bars show the standard error.

### Droplet pH, carbon dioxide partitioning and the rate of loss of infectivity at high RH

Replicating the physicochemical conditions that exist in the aerosol phase through bulk phase measurements is not possible except for conditions equivalent to the very highest RH. Under typical ambient conditions in the range 20-60% RH, solute concentrations are heavily supersaturated in equilibrated aerosols. In addition, the high surface-to-volume ratio in aerosol cannot be replicated, diminishing the potentially significant role of surface processes at the gas-liquid boundary and ignoring the influence of the rapid microphysical dynamics including the coupling of heat and mass transfer. However, certain elements of the airborne change in droplet composition can be replicated in the bulk phase by simulating the concentrations of various components in the droplet at concentrations equivalent to equilibration at high RH. The steady concentrations of solutes when the aerosol is equilibrated at 90% RH are approximately a factor of 10 higher than in the starting droplets at a water activity of 0.995 (Fig. S4), a concentration that can be replicated in the bulk. However, exposing SARS-CoV-2 to 10x MEM did not result in any observable loss of viral infectivity within 20- minutes (Fig. S6). This suggests that this increased concentration of culture media solutes is unable to account for the rate of loss of infectivity in the aerosol phase.

In addition to changes in the concentration of solutes that occur on equilibration to the ambient RH, it is possible that pH of aerosol droplets containing MEM can change rapidly. Although the sensitivity of SARS-CoV-2 infectivity both to high and to low pH has been reported (35, 61), these studies do not report measurements on a timescale relevant to the rapid loss of infectivity reported here in the aerosol phase. To investigate whether there is loss of infectivity with change in pH on a similar timescale to the loss of infectivity observed in the CELEBS measurements, SARS-CoV-2 was suspended in tissue culture media at varying pH for 20 minutes, before dilution into neutral media and plating onto cells for infectivity quantification (Fig. 5A). Although no significant decrease in infectivity was observed after 20 minutes at pH ranging from 5.6 – 9, the average infectivity was diminished considerably above pH 9.5 so that only 7% of the virus remained infectious after 20 minutes at pH 11.2. These bulk phase studies suggest that the pH would have to increase to around 11 to explain the deactivation observed in the aerosol phase at 90 % RH after 20 minutes. We therefore considered whether such high pH could be present in the aerosol droplets at high RH conditions in our experimental system.

**Figure 5.**
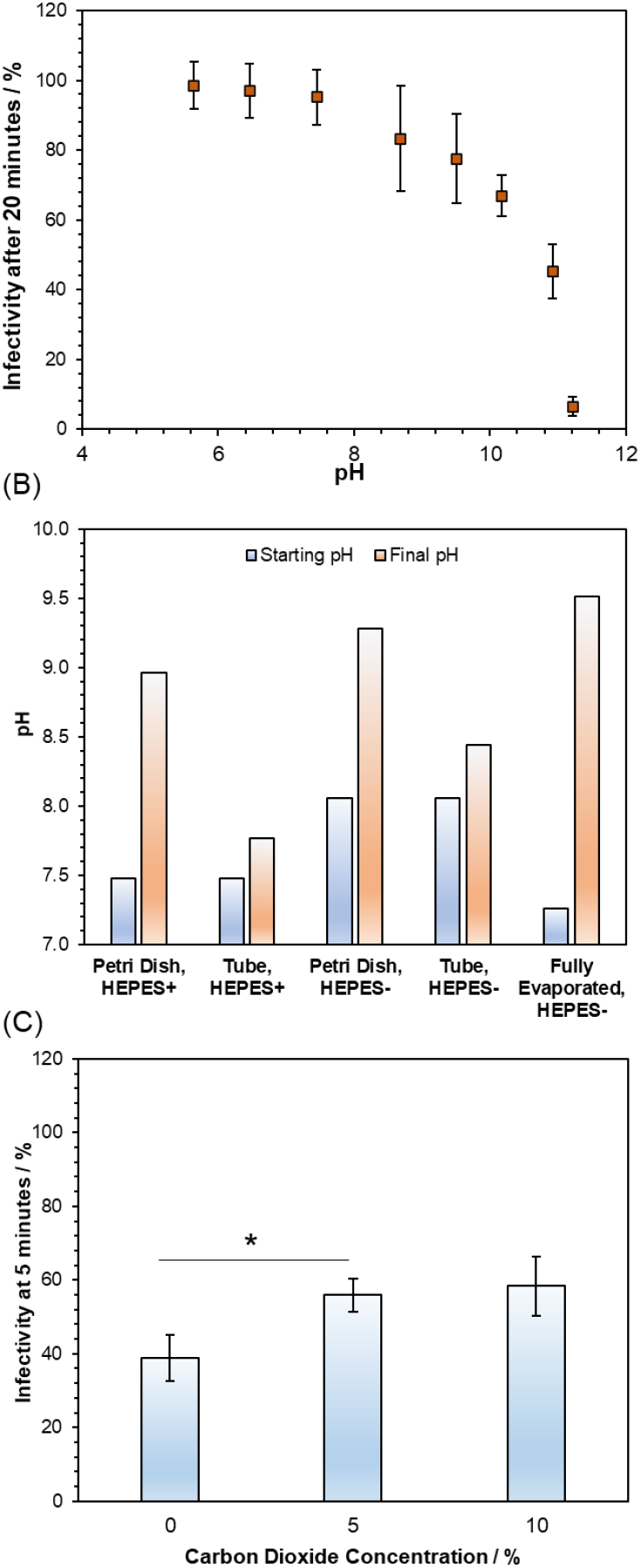
The Role of pH in SARS-CoV-2 Airborne Loss of Infectivity. (A) Bulk % infectivity of SARS-CoV-2 (REMRQ0001) after a 20-minute incubation in DMEM 2% FBS altered to a range of pHs, before being diluted back into neutral media and plated onto cells. Datapoints are the mean of ≥3 measurements with error bars showing the standard error. (B) The pH changes that tissue culture media (in this case DMEM) underwent when exposed to open air. DMEM was left in an open petri dish or 50 ml tube both with and without HEPES and the initial pH (blue bar) and the pH after 20 minutes (orange bar) was measured. The same measurement was carried out using thin layers of DMEM that were allowed to evaporate to 10% of their original volume over the course of 24 hours (labelled Fully Evaporated). (C) 5-minute levitations were carried out with SARS-CoV-2 (REMRQ0001) at 90% RH with varying CO_2_ concentrations mixed into the gas flow. Bars show the mean of 15 measurements for 0% CO_2_, 16 measurements for 5% CO_2_, and 6 measurements for 10% CO_2_ with the error bars showing the standard error. *p<0.03 between 0% and 5% CO_2_.

The equilibration between dissolved bicarbonate anions and gaseous CO_2_ is particularly important to consider for many respiratory secretions as well as the tissue culture media often used in experimental studies of airborne viral survival (62). A set of coupled equilibria is established with a bicarbonate concentration that responds to changes in the level of gas phase CO_2_, typically at elevated gas phase concentration for cell culture and 50,000 ppmv in exhaled air, specifically,

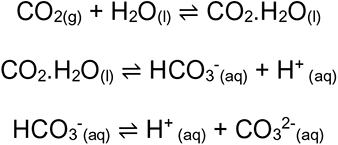

Cell culture media typically contain 20 - 50 mM bicarbonate (46, 47) to buffer the aqueous solution at a pH ∼7.4 when the CO_2_ is at an elevated concentration, 4-5% by volume. For bicarbonate in exhaled salivary aerosol (63), the lower gas phase CO_2_ concentration in the environment after exhalation (0.04%) results in a change in the equilibrium concentration of bicarbonate in the aerosol by shifting the equilibria towards CO_2_.H_2_O_(l)_ and eventually CO_2(g)_, leading to particle-to-gas phase partitioning of CO_2_. Indeed, for laboratory studies of airborne survival, the aerosol is often generated in an environment devoid of CO_2_, as is the case here, leading to irreversible evaporation of dissolved CO_2_ into the gas phase. As evaporation occurs, the available H^+^ concentration diminishes, and the pH can be expected to rise.

As a bulk analogue experiment, bulk tissue culture medium was exposed to ambient air for an extended time period (Fig. 5B). After 20 minutes in an open Petri dish, the pH of DMEM (formulated with the same concentration of bicarbonate as the MEM used in the levitations) rises from 8 to 9.3. Adding HEPES reduces the initial pH of the medium, but the pH was still found to increase significantly after exposure to air, increasing from 7.4 to 9. When the DMEM solution was kept in an open 50 ml tube rather than a petri dish, decreasing the surface area for interaction, the rate of the rise in pH also decreased. A final experiment was carried out in which thin layers of DMEM were placed in petri dishes and allowed to evaporate to 10% of the starting volume over the course of 24 hours, replicating both the CO_2_ and H_2_O equilibration that takes place in airborne droplets. The combination of CO_2_ equilibration and volume loss resulted in the greatest pH rise, increasing from 7.25 to 9.5. The particle-gas partitioning can be expected to occur more rapidly than in any of these bulk examples because of facile transport across a droplet surface with high surface-to-volume ratio. Whilst the presence of bicarbonate buffers biological fluids such as saliva (63) when they are in the respiratory tract (or in a CO_2_ supplied incubator), the decrease in the concentration of dissolved bicarbonate through the irreversible loss of CO_2_ following their aerosolisation will cause droplets to become more alkaline.

Sodium bicarbonate accounts for ∼20% of the solute mass in MEM with ∼65% sodium chloride by mass. With the loss of bicarbonate from MEM solution droplets, through irreversible evaporation of CO_2_, the reduction in solute mass should lead to a reduction in the wet equilibrated size of the droplet with less solute able to sustain less water in the condensed phase. Indeed, it was possible to observe a long-time slow loss of CO_2_ and dissolved solute using the CK-EDB for MEM solution droplets (Fig. S7) and for mixtures of NaCl and NaHCO_3_ (Fig. S8), the two dominant salts. Droplets of both MEM and sodium bicarbonate continue to decrease in size for longer than the time required for the water activity to equilibrate to the gas phase RH. Indeed, the vapour pressures inferred from the data in Fig. S8 with varying RH (0.0092, 0.014 and 0.052 Pa at 60, 75 and 90 % RH) are consistent with calculations using the E-AIM model (http://www.aim.env.uea.ac.uk/aim/aim.php) (64) for the vapour pressure of CO_2_ above supersaturated carbonate solutions at the same water activities (∼0.01 Pa and increasing with increase in RH). By contrast, the vapour pressure of CO_2_ from bicarbonate solutions at the same RHs are considerably higher (∼100 kPa) and the particle-gas partitioning can be expected to occur extremely rapidly in << 1 s following aerosol droplet exhalation or generation, a process that can be expected to already be completed by the time the aerosol droplets are captured by CELEBS or the CK-EDB.

During the evaporation of water, as the moisture content of the aerosol equilibrates, the solutes surpass solubility limits for various salts. The initial water activity of the starting droplets can be estimated as 0.9952 by considering the dominant ionic species alone (Na^+^, Ca^2+^, Cl^-^ and HCO_3_^-^) using the E-AIM model (64). Calcium carbonate is particularly insoluble and becomes supersaturated from very early on in the evaporation process, successively followed by other binary and mixed salts, specifically CaNa_2_(CO_3_)_2_.5H_2_O(s), Na_2_Ca(CO_3_)_2_.2H_2_O(s), NaHCO_3_(s) and finally NaCl(s) as water activity decreases. The droplet becomes saturated with respect to the first two salts above a water activity of 0.9, sodium bicarbonate at ∼ 0.9 and NaCl below 0.8. Indeed, we observe the precipitation of salts during the droplet equilibration process as the water activity transitions through to the final equilibrated value, with significant supersaturation required for each before crystallisation occurs (see Fig. 2B). Until the crystallisation of NaCl(s), which only occurs at the very lowest RHs of 50% and below, a partially deliquesced particle containing crystalline inclusions along with an aqueous phase, leads to considerable variability in the remaining infectivity of the virus (Fig. 1B).

It can be hypothesised that increasing the concentration of CO_2(g)_ around the droplet would reduce the irreversible loss of bicarbonate from the droplet and could mitigate a pH-driven loss of infectivity. CO_2(g)_ was added to the airflow during CELEBS levitations at high RH and the infectivity of SARS-CoV-2 measured after 5 minutes (Fig. 5C). The elevation to a gas phase concentration of 5% by volume CO_2_ (equivalent to 50,000 ppmv) around the droplet at 90% RH results in a small but significant increase in the remaining infectivity of SARS-CoV-2 after 5-minutes when compared to ambient CO_2(g)_ (0.04%). Increasing the steady CO_2(g)_ concentration around the trapped droplet cannot mitigate loss of infectivity from pH changes during the initial travel of the droplet to the trapping region. Whilst it is possible that elevated CO_2(g)_ around the droplet may have other physicochemical effects on the droplet in addition to decreasing the pH, this measurement provides further evidence that increased droplet pH is at least partly responsible for the observed falls in viral infectivity at high RH.

The influence of pH on infectivity is expected to be relevant in respiratory aerosols as the underlying physicochemical properties of exhaled aerosol (saliva) and MEM are similar, and numerous studies have demonstrated that exhaled breath condensate is alkaline (48, 65–67). This dynamic is in stark contrast to environmental aerosols such as sea spray, where following generation the pH of the sea spray, droplets will become more acidic through the uptake of acidic gases such as HCl, and SO_x_ (68). Exhaled aerosol is generated in an environment with an extremely high concentration of an acidic gas (4-5% by volume CO_2_) that can only be reduced once exhaled. This contrast in pH behaviour following generation is clear when comparing studies of collected sea spray pH (68, 69) with those of collected exhaled breath condensate (48, 66, 67, 70). In short, while the vast majority of ambient aerosol may be acidic, exhaled aerosol can be expected to be alkaline. The pH of exhaled and model respiratory aerosols is an area in need of further study, with a need for measurements across a broad range of time scales, droplet compositions (saliva, sputum, MEM, DMEM), and environmental conditions (RH, [CO_2(g)_]).

### Comparison to rotating drum studies of SARS-CoV-2

We compare the current measurements with these previous studies in Fig. 6. A motivation of our combined approach using CELEBS and CK-EDB is to identify the fundamental physicochemical parameters that dictate viral infectivity in the aerosol phase; to go beyond general associations, such as those between RH and infectivity, and address the more challenging and informative questions allowing the identification of mechanistic causation rather than just correlation. By taking this approach, it has been shown that SARS-CoV-2 undergoes a rapid deactivation in the first few minutes following droplet generation, and that this deactivation is driven by efflorescence at low RH and possibly by an increase in droplet pH at high RH resulting from irreversible partitioning of CO_2_ into the gas phase. Throughout the pandemic there have been several reports of the aerostability of SARS-CoV-2 using the Goldberg rotating drum (12–15). However, given the relatively short timescale over which the majority of this deactivation occurs which drum experiments cannot observe, and the importance of the physicochemical properties of the droplet in driving the deactivation, it is unsurprising that data collected from rotating drums report a longer lifetime for the virus in the aerosol phase.

**Figure 6.**
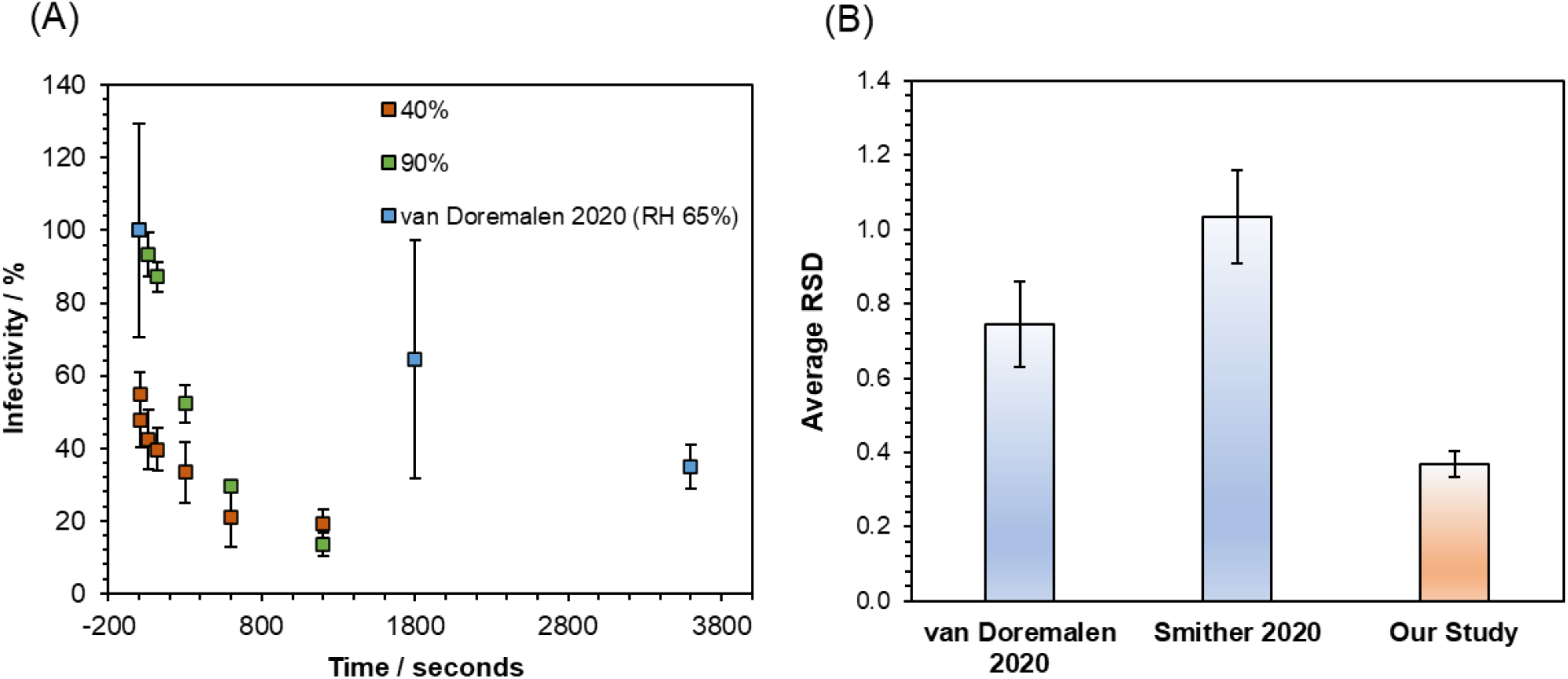
Comparison of CELEBS measurements to rotating drum measurements of SARS-CoV-2. (A) Data from Figure 1a plotted alongside the SARS-CoV-2 airborne stability curve published by van Doremalen et al. Data from van Doremalen was normalised to their earliest measurement to allow direct comparison to the CELEBS data. CELEBS data plotted in orange and green is as described in Figure 1. Data from van Doremalen et al is plotted in blue with datapoints showing the mean of 3 measurements each and error bars showing the standard error. (B) Bar chart showing the RSDs calculated from SARS-CoV-2 aerostability datasets published by van Doremalen et al, Smither et al, and from this study. RSDs were calculated by dividing the standard deviation by the mean for each datapoint published. The average and standard error of these RSDs was then calculated and plotted. RSDs are the mean of 5 RSDs from van Doremalen 2020, 17 RSDs from Smither 2020, and 19 RSDs from our study.

Rotating drums have a poorly defined *time-zero*, meaning that the benchmark infectivity to which later time data is compared is poorly defined. The number of droplets suspended, their initial size, and viral units per droplet are both variable and uncontrolled, and thus must be inferred offline. RH profiles, while they appear to be commonly collected in rotating drum experiments, are rarely reported in their entirety, with many drum studies only reporting a single RH value for each measurement (12, 14). The location of the RH probe within the experiment, and whether the value is taken at a particular time or is an average across their experiment, is not always reported. Regardless, the RH recorded by a probe is likely unrelated to the RH trajectory that an aerosol droplet experiences as it passes from the nebulisation source into the rotating vessel. Indeed, the nebulisation of a cloud of aerosol at high number concentration likely leads to some buffering of the RH in the gas phase, sustaining a higher value that would be typical of the very low respiratory aerosol concentrations actually generated (20). These uncertainties may make the influence of processes such as efflorescence on infectivity challenging to infer.

Comparisons of the time dependence and precision of the CELEBS measurements to those from rotating drum studies are reported in Fig. 6. The time-resolution of the drum measurements make the initial decrease in infectivity challenging to identify. Indeed, the times of the indicated points (Fig. 6A) should not be taken as the time-resolution, for the reasons discussed above. In addition, the average relative standard deviation (RSD) from the CELEBS measurements is 0.37 (Fig. 6B), compared to 0.66 from van Doremalen et al. (12) and 1.03 from Smither et al. (13) Two further papers do not report sufficient information to estimate RSDs (14, 15). The smaller RSD from the CELEBS is likely the result of the more stable environmental conditions, a more reproducible monodisperse droplet generation process, and improved methodology for viral infectivity quantification (37). Furthermore, CELEBS experiments are more straightforward to perform, allowing for more repeat measurements for each condition and leading to a high degree of confidence in the mean % infectivity values reported.

The nebulization of bicarbonate buffered solutions into a confined volume results in the elevation of the CO_2_ gas concentration (Fig. S9). The magnitude of this elevation is dependent on many variables, including the pH of the nebulized solution, the nebulization time, and the drum volume. A survey of the literature failed to identify a single article where the CO_2_ levels within a rotating drum was reported. As reported (Fig. 4D), CO_2_ in the gas phase reduces the degradation of the virus likely by limiting the rise in droplet pH. CO_2_ cannot be removed selectively during a rotating drum study and the conditions likely support greater SARS-CoV-2 longevity. Accumulation of CO_2_ is not an issue in CELEBS due to the constant flow of compositionally controlled air being maintained over the trapped droplets. In addition to potential issues with the pH of the airborne droplets in the rotating drum, it is also possible that the pH of the solution within the nebulizer may increase during the nebulization process (Fig. S10), directly affecting the viral infectivity prior to nebulization (Fig. 4A).

## Discussion

A combination of measurement strategies to probe the changes in airborne viral infectivity with time and the physicochemical transformation dynamics of the host aerosol is crucial to improve our understanding of the influence of environmental (such as RH, temperature) and biological (such as spike protein mutations) parameters on the transmission of viruses in the aerosol phase. While the current general consensus is that the half-life of SARS-CoV-2 in the aerosol phase is between 1-2 hours, if not longer, we report an initial rapid decline in infectivity within a few seconds to minutes of aerosol generation. Under all conditions measured, the majority of SARS-CoV-2 is inactivated within 10 minutes of aerosolization. Further research is required to determine for how long the remaining fraction persists and how this may depend on the viral load in the aerosol. The high-time resolution infectivity measurements reported here are uniquely accessible to the CELEBS technology and can only be understood once the detailed aerosol microphysics are fully explored. Although we do not report measurements in artificial or real saliva, the culture media used do have many of the same characteristics of real respiratory secretions, particularly the high concentration of inorganic ions that dominate the phase behaviour and water content of the aerosol, along with bicarbonate ions that partition CO_2_ into the gas phase on aerosolisation. In addition, the initial water activity of the aerosol is consistent with the high RH of the respiratory tract and the aerosol generation process generates isolated droplets that must respond rapidly to the surrounding environmental conditions, typical of the very low concentrations of aerosol exhaled in infected individuals.

The aerostability data reported here are consistent with a view that the virus is primarily spread over short distances. Often, the assumption is that short distance transmission is caused by large droplets that fall to the ground more quickly and therefore do not travel as far. The rapid loss of infectivity demonstrated in these measurements provides an alternative explanation for a short transmission distance, with rapid airborne losses of viral infectivity making transmission decreasingly likely as distance from the particle source is increased, even if the particles that contain the virus are small and able to travel long distances. This loss in infectivity is compounded by the considerable dilution in aerosol concentration that results following exhalation and transport beyond the short range.

We do not observe the characteristic “V-shape” relationship between RH and virus stability, where maximum virus loss occurs around RH = 50%. Rather, the largest loss of infectivity was observed at the lowest RHs. Previously Goldberg drum studies have not identified a strong dependence for SARS-CoV-2 survival on RH (13). However, following the initial loss of infectivity, the virus within the now dry particle appears to be somewhat stable when compared to the higher RH. Thus, if the initial rapid decrease in infectivity is not accounted for when reporting RH stability data, a “V-shape” relationship may be identifiable. However, not accounting for changes in viral infectivity that take place immediately after particle generation prevents the accurate coupling of airborne stability measurements with measurements of initial virus shedding, limiting the value of the V-shape relationship.

The rapid loss of SARS-CoV-2 infectivity through droplet efflorescence at RH < 45% suggests that dry air may help to limit overall exposure. However, investigation of the impact that lowering RH has on particle transport in the exhalation jet is required to confirm this. The large impact of efflorescence on SARS-CoV-2 infectivity, indicates that measuring the impact of environmental conditions on phase change in respiratory secretion aerosols may provide useful insights into COVID-19 transmission. Further research is needed to confirm with more certainty the degree to which pH is involved in the airborne loss of SARS-CoV-2 infectivity at high RH and to determine the exact mechanism by which the pH rise is deactivating the virus. The importance of elucidating of the role of pH in the survival of SARS-CoV-2 in the aerosol phase cannot be understated. A literature survey found no manuscripts indicating that the alkaline nature of exhaled aerosol may affect viral infectivity. Contrarily, it has been reported that viruses may be inactivated by acid in the aerosol phase (71).

Elevation of CO_2_ levels within a room is taken as a clear sign of occupancy and poor ventilation. There has been increasing discussion surrounding the use of CO_2_ monitors as a means of determining the relative risk of COVID-19 transmission in various settings. The data from this study give further credence to this approach. Not only is elevated CO_2_ an indication of a densely occupied, poorly ventilated space, but it could also be indicative of an environment in which SARS-CoV-2 is more stable in the air. The precise elevation in CO_2_ required for an observable improvement in SARS-CoV-2 is unknown and requires further investigation (5% CO_2_ is not a concentration reached in typical indoor environments), but it is possible that this is an additional risk presented by poorly ventilated, densely occupied settings. If so, CO_2_ monitors may present an immensely valuable means of assessing the relative risk of different indoor environments.

The approach taken here has clearly demonstrated the value of a combined approach that considers both the aerosol microphysics and biological processes in tandem, demonstrating that underlying parameters that drive SARS-CoV-2 inactivation in the aerosol phase are particle phase and pH. In further research, we intend to explore these processes over an even wider range of times, conditions, and virus variants. There also remain unanswered questions as to exactly how phase change and high pH deactivate the virus. Do these processes rupture the viral envelope, or impart an irreversible modification to the spike protein? Is the effect of pH the result of direct deprotonation of viral molecules, or is it an indirect effect caused by alterations to the solubility of other components within the droplet? Answering such questions would provide key insights into the physicochemical and biomolecular processes governing SARS-CoV-2 transmission, and airborne pathogen transmission more broadly. It is only by pushing the limits of aerobiology to this deeper level, that we can hope to understand how best to prevent the airborne spread of disease.

## Materials and Methods

Details of virus strains, and methodologies for: virus and cell culture, viral infectivity quantification, bulk stability measurements, CK-EDB measurements, and falling droplet column measurements found in SI Materials and Methods.

### Generation and Trapping of Droplets

The reservoir of a droplet-on-demand dispenser (MicroFab) is filled with MEM 2% FBS. The application of a square waveform to the piezoelectric crystal results in a compression wave that passes through the dispenser’s orifice and initiates the formation of a jet which forms droplets of uniform size with each pulse. A DC voltage is applied to an induction electrode, positioned 2-3 mm from the dispenser tip, which leads to an ion imbalance in the jet, resulting in a droplet with a net charge. The presence of this net charge interacting with the electrodynamic field of the CELEBS/CK- EDB leads to confinement of the droplet within the null field point.

### CELEBS Airborne Longevity Measurements

The environmental conditions were set by adjusting the Peltier voltage and polarity to set the temperature and the ratio of dry to wet air to set the humidity. SARS-CoV-2 suspension is drawn into a 1 ml syringe which is then attached to the instrument and used to feed the virus solution to the droplet dispenser via a remotely operated motor. Droplets are then generated and trapped as described above. Once the desired time is reached, an isolation plate is retracted causing the electric field to be set to zero; then, the droplets are pulled down into a plate containing 5-10 ml of DMEM 2% FBS so that the remaining virus can be quantified (see extended methods in SI). For each measurement, two levitations are carried out. First a short levitation of <5 seconds is used to measure the initial virus per droplet number, and then a second levitation for which the droplets are kept in the trap for the length of time being investigated.

## Data Availability

All data produced in the present study are available upon reasonable request to the authors

## Acknowledgements

This work was funded by the NIHR-UKRI rapid COVID-19 call, the Elizabeth Blackwell Institute for Health Research, the University of Bristol, and the Medical Research Council. Additionally, this work was supported by funding from the PROTECT COVID-19 National Core Study on transmission and environment, managed by the Health and Safety Executive on behalf of HM Government. AH and MOF received funding from the BBSRC, project BB/T011688/1. HO is supported by funding from DSTL and EPSRC. The authors would also like to thank Robert Alexander for his contributions to discussions on aerosol pH.

## Extended Materials and Methods

### Virus Strains and Culture Methods

Vero E6 cells modified to constitutively express TMPRSS2 (Vero E6/TMPRSS2 cells (72); obtained from NIBSC, UK) were cultured at 37°C and 5% CO_2_ in Dulbecco’s Modified Eagle Medium (DMEM, high glucose; Sigma, UK) supplemented with 10% foetal bovine serum (FBS, Sigma), 100 units/ml penicillin (Gibco, UK), 100 µg/ml streptomycin (Gibco, UK), and L-glutamine (Gibco, UK). The SARS-CoV-2 viral isolates, SARS-CoV-2/human/Liverpool/REMRQ0001/2020 (REMRQ0001, GenBank: MW041156.1), hCoV-19/England/02/2020 (GISAID ID: EPI_ISL_407073) and the “Bristol” variant derived from it (BriSΔ) in which spike amino acids 679-687 (NSPRRARSV) had been deleted and replaced with Ile (59) were isolated as previously described (60). The SARS-CoV-2 variant hCoV- 19/England/204690005/2020 (lineage B.1.1.7 - Alpha variant; GISAID ID: EPI_ISL_693401) was kindly provided by Professor Wendy Barclay, Imperial College, London and Professor Maria Zambon, Public Health England. Stocks of SARS-CoV-2 isolates were produced by inoculation of Vero E6/TMPRSS2 cells at a multiplicity of infection (MOI) of 0.01 and incubating the cells for 48- 72 h in Eagle’s minimum essential medium plus GlutaMAX (MEM, Gibco, ThermoFisher, cat# 41090036) supplemented with 2% v/v FBS and 0.1 mM non-essential amino acids (MEM 2% FBS). The culture supernatants were clarified by filtration through a 0.2 µM filter and stored in aliquots at − 80 °C. The titre of the stocks was determined by preparing 10-fold serial dilutions in MEM 2% FBS which were added to 1 × 10^4^ Vero E6 cells in the same medium in each of 12 wells of a 96-well plate. Plates were incubated at 37 °C for 4 - 7 days and then examined for cytopathic effect (CPE). The TCID_50_ was calculated according to the method of Reed and Muench (73).

### Quantification of Virus

SARS-CoV-2 in aerosol experiments was quantified by measuring the occurrence of viral induced CPE on Vero E6/TMPRSS2 cells seeded in 96-well plates. Viral stock titres were calculated by TCID_50_. A 10-fold dilution series of the virus suspension was prepared in DMEM 2% FBS and each dilution was then used to infect a row of cells with 100µl of virus. The plate was incubated for 4 - 6 days at 37°C 5% CO_2_ and CPE then observed by microscopy. By counting the number of infected wells on each row the virus titre can be calculated using the Reed-Muench method (73).

When the virus concentration was lower than 10^2^ infectious units ml^-1^, as was the case for levitated virus, an alternative approach was used. The entire neat suspension was used to inoculate Vero E6/TMPRSS2 cell seeded wells in a 96 well plate with 100µl of sample per well. Depending on the volume of the sample, the number of wells inoculated would vary, but typically the SARS-CoV-2 levitations would be deposited into 6 ml of media and then used to inoculate 60 wells in a plate, such that the outermost wells of the plate were left empty. The plate was then incubated for 4 - 6 days at 37°C and the cytopathy assessed by microscopy. To calculate the amount of virus in the plate, a rearranged form of the Poisson distribution equation was used:

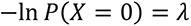

Where *P(X=0)* is the proportion of uninfected wells (calculated by dividing the total number of uninfected wells by the total number of inoculated wells) and λ is the calculated infectious units per 100µl. By multiplying λ by the total volume of the sample and then dividing it by the number of droplets deposited into the sample, the number of infectious units per droplet can be calculated.

### Bulk Stability Measurements

To assess the stability of SARS-CoV-2 in bulk solutions, the virus stock was first diluted to 10^-4^ in the test solution (DMEM or MEM with altered pH or solute concentration). This 10^-4^ solution was then incubated in a sealed tube at room temperature for 20 minutes. 100 µl was taken from this and diluted back into 900 µl of normal DMEM 2% FBS. 500 µl was then added to 19.5 ml of DMEM 2% FBS (when inoculating 3 sets of 60 wells). This final dilution (a concentration of 10^-6.3^ of the original stock) was then used to infect cells for the quantification of remaining viral infectivity by the same method used to quantify virus post levitation. This 10^-6.3^ dilution was chosen as the SARS-CoV-2 stocks typically have a TCID_50_ of around 10^7^ ml^-1^ meaning that a 10^-6.3^ dilution would result in around 50% of wells being infected granting accurate viral quantification. This dilution can be adjusted for different stock concentrations. A portion of the stock was also immediately diluted to 10^-6.3^ and used to infect cells to calculate a T0 control value to which the incubated sample can be normalised.

### CK-EDB Measurements of MEM

The droplets were generated and trapped as described above. Once confined, approximately 100 ms after droplet formation, the droplet was illuminated by a 532 nm laser (Laser Quantum, Ventus continuous wave [CW]). A nitrogen gas flow of 200 mL/min at a temperature of 20 °C and a set RH (range from ∼0% to >90%) was passed directly over the droplet. As the droplet changes size, the electrodynamic field was manipulated to account for these changes and ensure that it remained confined within the centre of both the trap and laser beam.

A CCD camera (Thorlabs) collected the light scattered from the droplet in the near-forward direction at a central scattering angle of 45°. Images of the phase function, with an angular range from 32° to 58°, were collected every ∼10 ms providing high time resolution measurements of particle size and morphology. This range was selected for two reasons: it is readily accessible to numerous other individual droplet analysis devices and the central viewing angle of 45° allowed access to the region of the near-forward scattering that corresponds to the range of applicability of the geometric optics approximation (up to 60°) for particle sizing.(74)

When a droplet was spherical and homogeneous, its radius and refractive index of the droplet could be estimated (using a prescribed relationship between droplet radius and refractive index(75)) by fitting a collection of time dependent phase functions with a library of Mie theory simulations (as seen in Figures 3b and 4b). This method is computationally demanding. Alternatively, the absolute radius of the droplet could be inferred from the average angular difference between the maxima within the phase function using the geometrical optics approximation.(74) This approach allowed for rapid analysis of each collected frame in real-time.

### Falling Droplet Column Imaging of MEM

See Hardy *et al* 2021 (58) for detailed methodology. Droplets of MEM 2% FBS were dispensed into an enclosed columnar chamber maintained at a constant RH by a flow of temperature and humidity-controlled air. The in-flight images were collected by a CCD camera with droplets illuminated by stroboscopic lighting from a high-power LED. Particles were collected at the bottom of the column and imaged by SEM.

---

**Figure S1.**
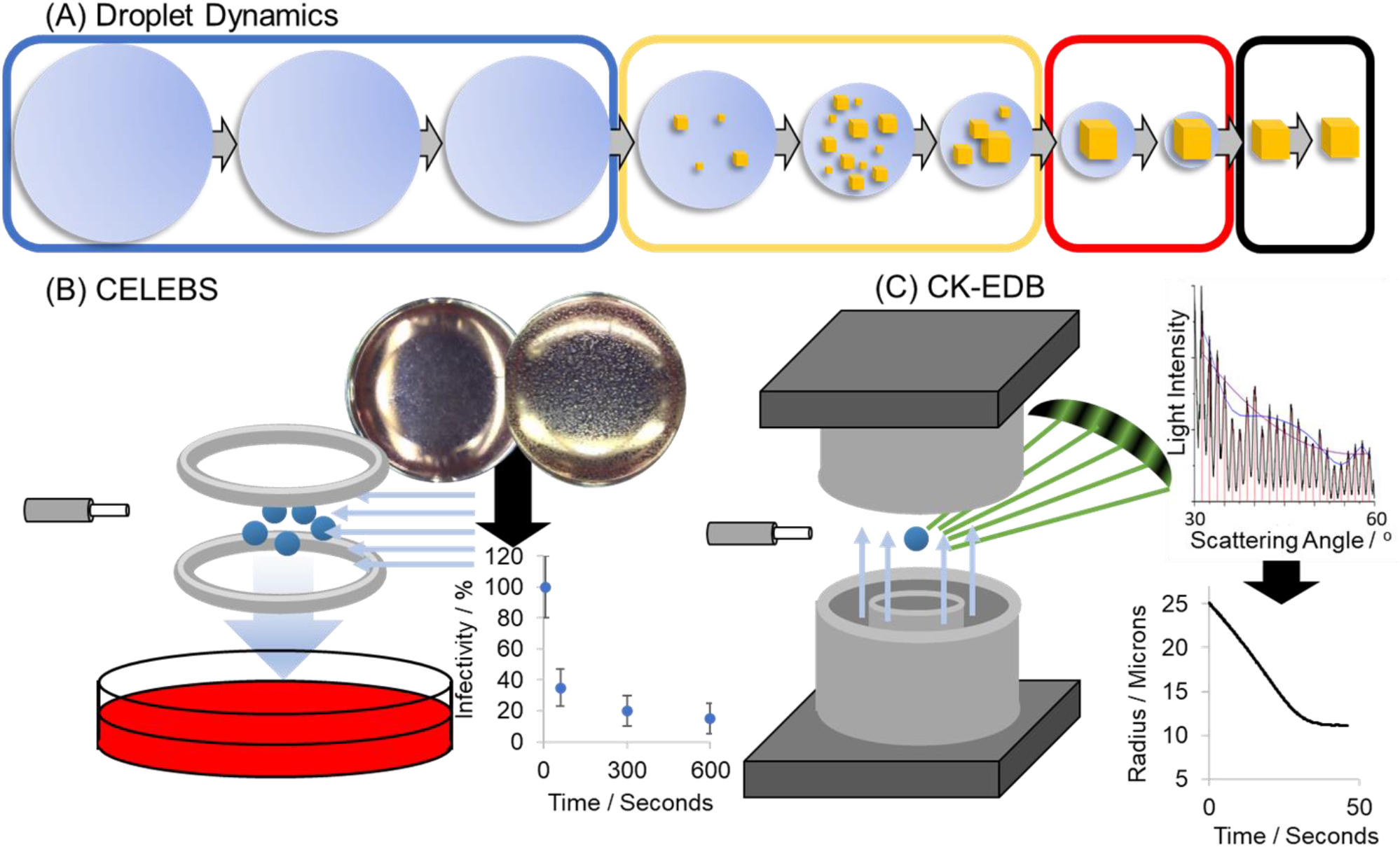
Experimental Approach. (A) Schematic of the physical changes that take place when an airborne aqueous droplet equilibrates to the surrounding relative humidity. On the left (blue box) the particle is an aqueous homogenous sphere. As the particle equilibrates to a sub-saturated RH, low-solubility solutes can precipitate inclusions within the droplet (yellow box). At a sufficiently low RH, the dominant solute (NaCl) can crystallise causing the particle to effloresce (black box). (B) Schematic of the CELEBS technique. Virus containing particles are levitated under controlled conditions and then deposited into media which is then plated onto a cell culture. By enumerating the cytopathic effect from that deposition, the amount of virus present can be quantified. (C) Schematic of the CK-EDB technique. Particles generated by the same droplet dispensers used in CELEBS are levitated under controlled conditions in the path of a laser. The physical changes that take place in that particle are studied through analysis of the light scattered by that particle.

**Figure S2.**
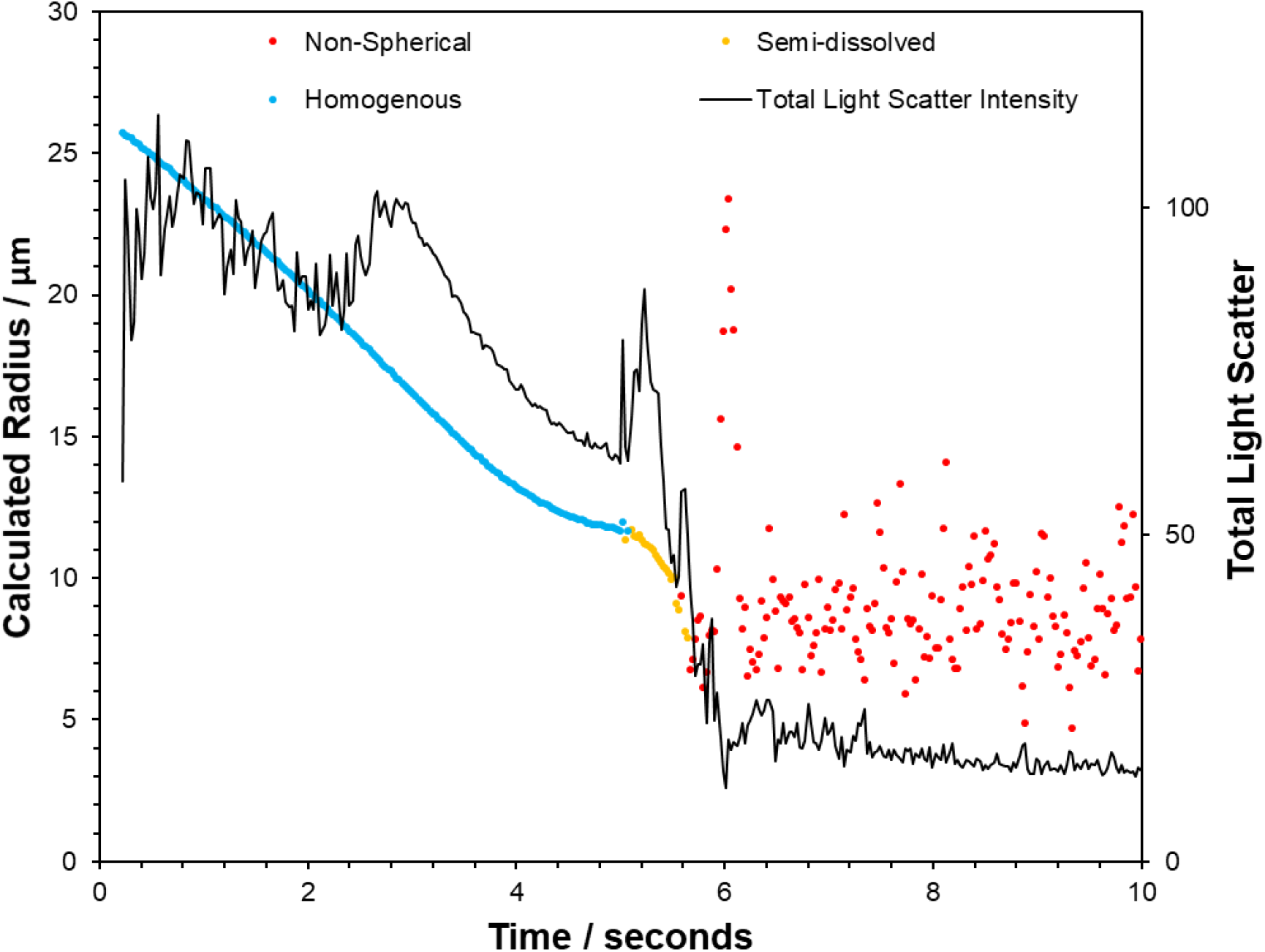
Sodium Chloride Efflorescence. A droplet of sodium chloride was trapped in the CK- EDB at 30% relative humidity (RH) and allowed to effloresce. The radius (plotted against the left y-axis) and structure was measured using the Mie scatter and is plotted in the coloured points. Blue points indicate a spherical homogenous droplet, yellow points indicate inclusions, and red points indicate a non-spherical particle (note that accurate sizing is not possible for non-spherical particles). The total light scatter intensity is plotted against the right-hand y-axis as a black line. The abrupt drop in total light scatter coincides with the efflorescence of the droplet.

**Figure S3.**
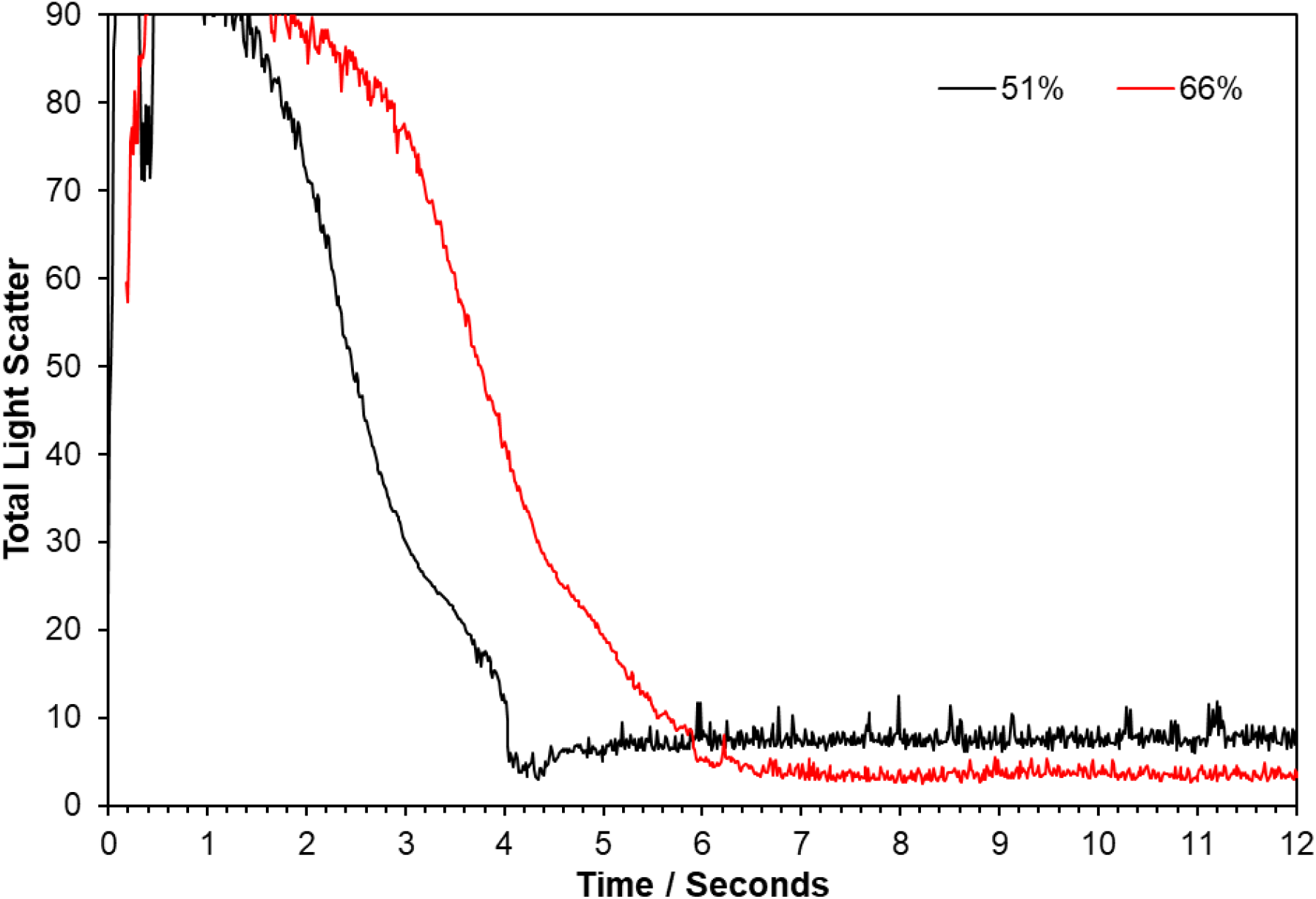
MEM 2% FBS Efflorescence. The total light scatter intensity was measured in the CK- EDB for droplets of MEM 2% FBS at a range of relative humidities. Shown here are the measurements at 51% RH (black line) and 66% RH (red line). At 51% RH the sudden drop in light intensity characteristic of efflorescence is observed. At 66% RH no efflorescence is observed.

**Figure S4.**
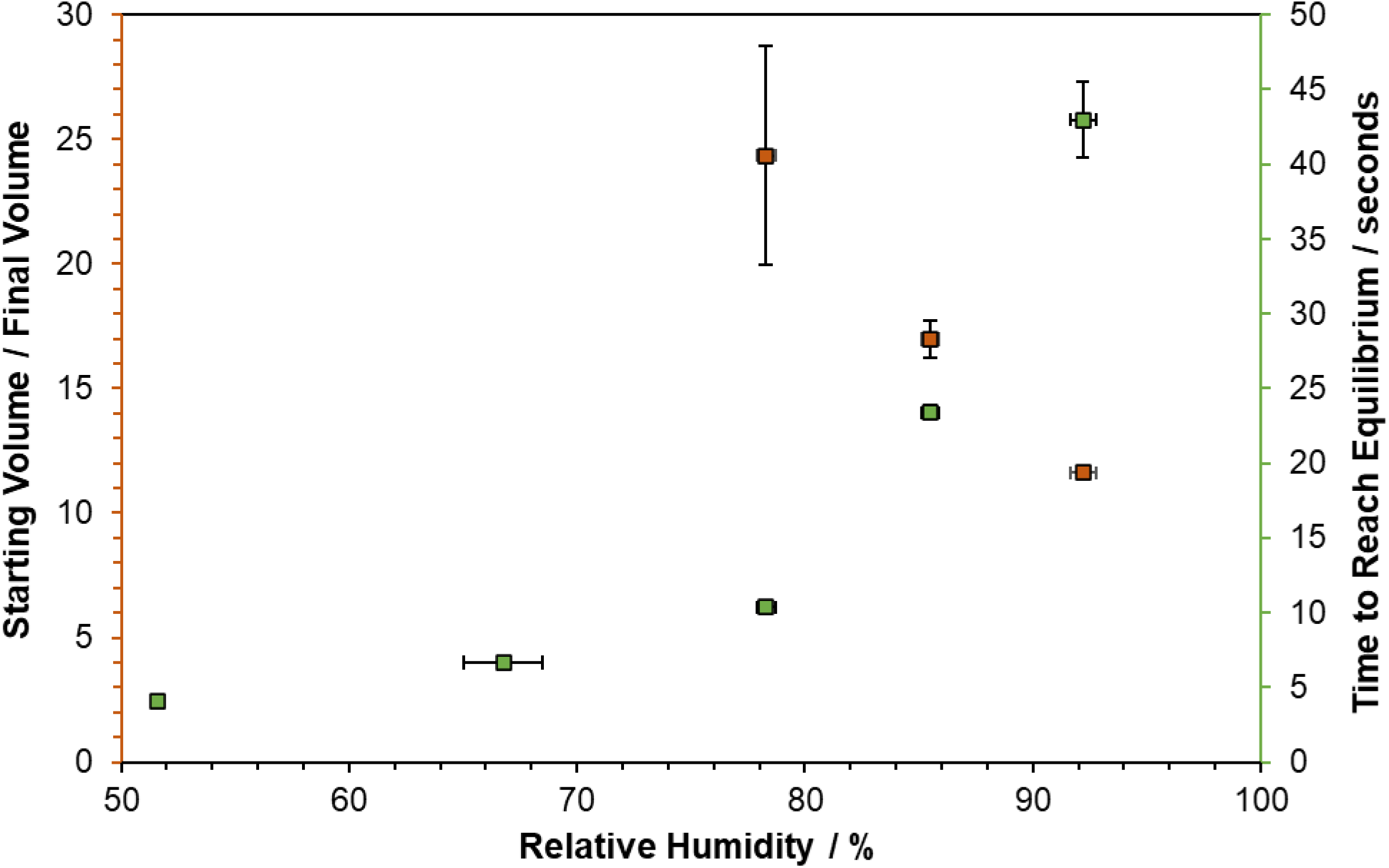
MEM 2% FBS Airborne Size Change. The ratio of initial volume of the droplet to the final volume droplet (orange, left-hand y-axis) and the time taken for the droplet to equilibrate (green, right-hand y-axis). Datapoints are the mean of 10 measurements. Error bars show the standard deviation of the RH (x error bars) and measurements (y error bars).

**Figure S5.**
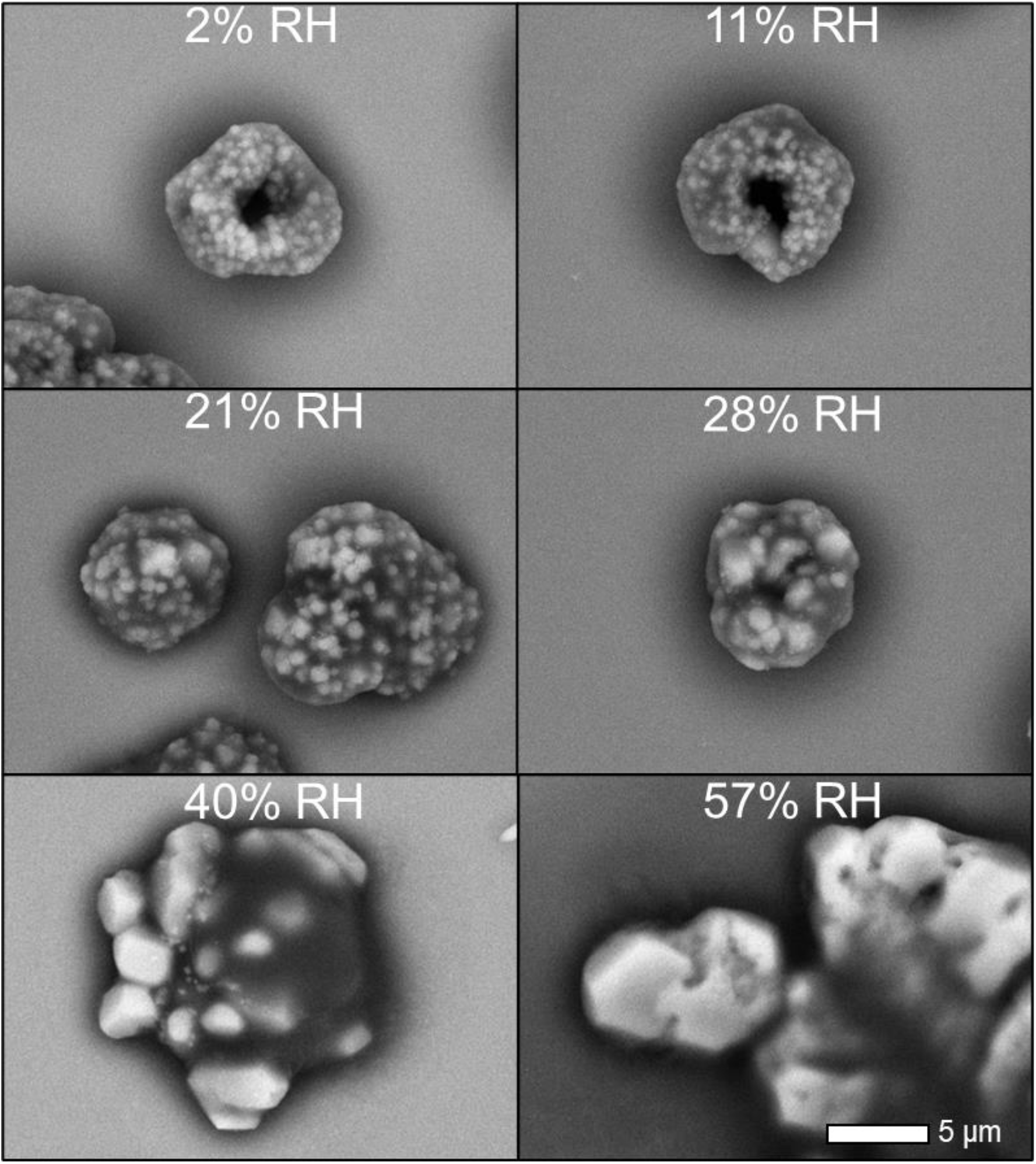
MEM 2% FBS SEM Images. SEM micrographs of MEM 2% FBS particles collected from the bottom of the falling droplet column after falling at a range of RHs. Note that the more spread-out salt crystals at 57% RH are likely the result of the particles still being liquid upon deposition. The scale bars (white at the bottom) show 5 µm.

**Figure S6.**
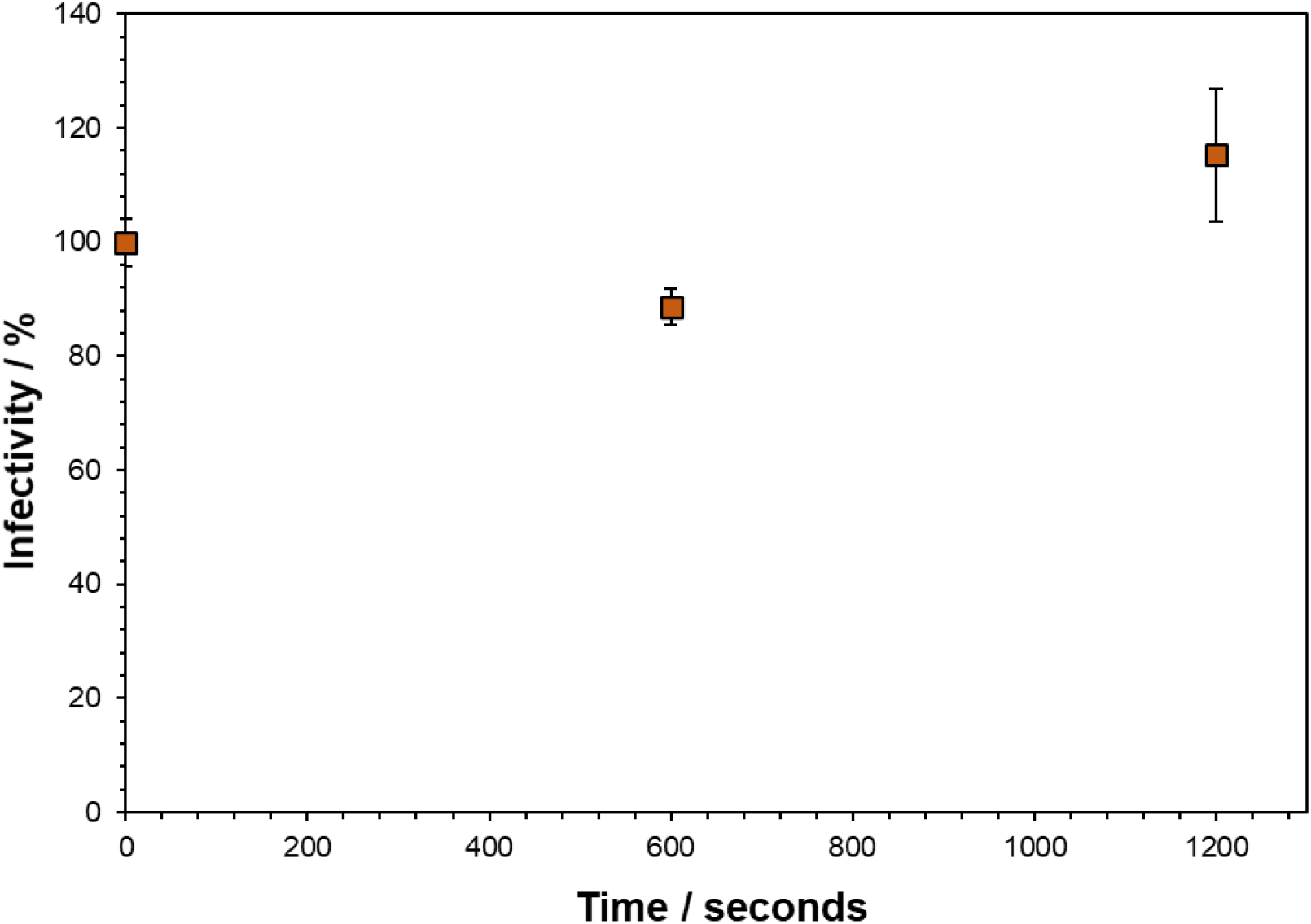
Bulk Survival of SARS-CoV-2 in 10x MEM. Bulk % infectivity measurement of SARS-CoV-2 incubated for 20 minutes in 10x MEM 2% FBS before being diluted back into normal media (DMEM 10% FBS) and plated onto cells. Datapoints are the mean of 6 measurements for 10 minutes and 3 measurements for 20 minutes with error bars showing the standard error.

**Figure S7.**
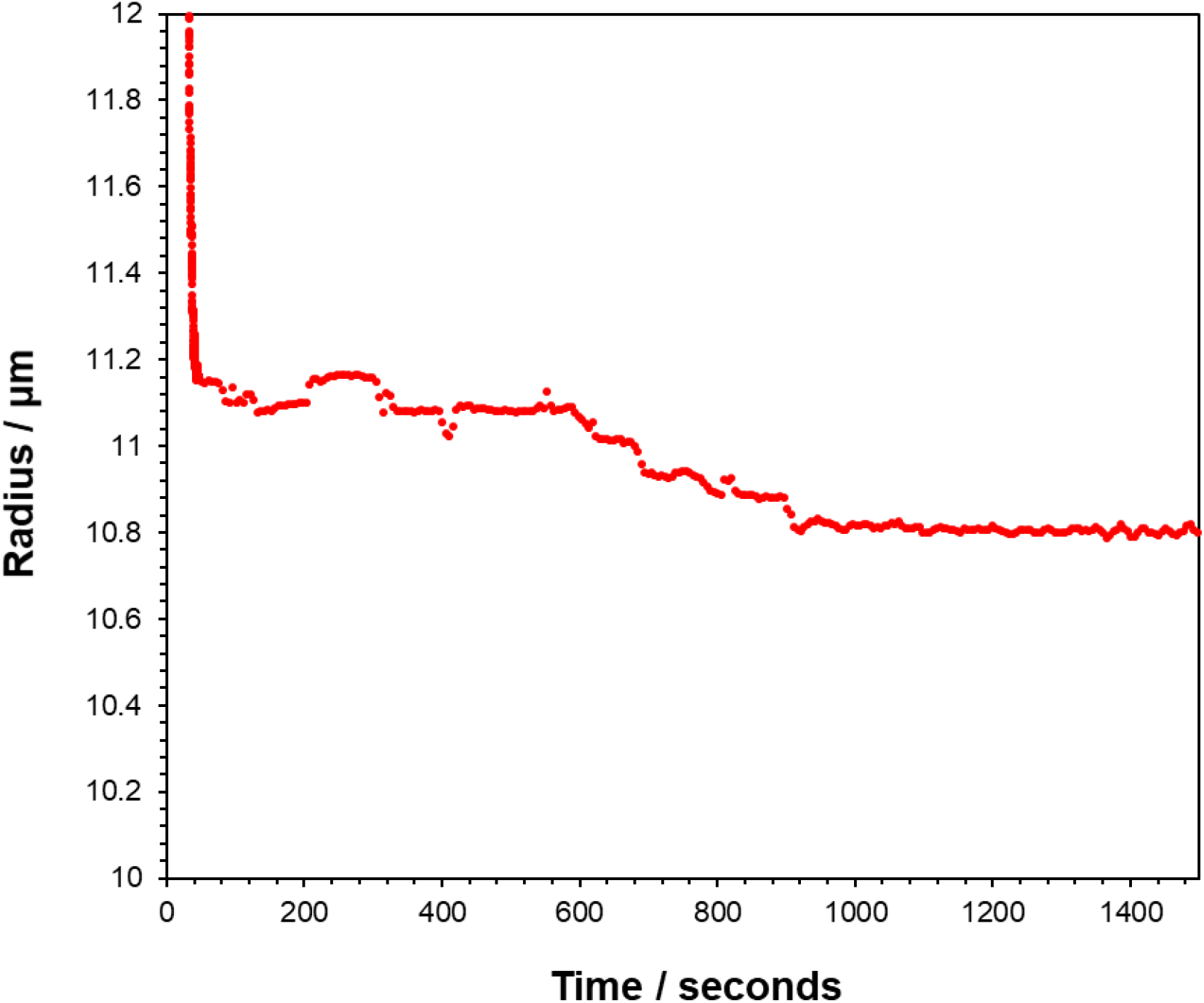
Change in DMEM droplet radius as a result of CO_2_ loss. CK-EDB measurement of an airborne MEM droplets shows the size continuing to decrease after the water equilibration is complete. The initial droplet radius was ∼25 µm and the RH was ∼90%. The initial size loss of the droplet finished after ∼55 seconds as expected for a culture media levitation at high RH. However, the size of the droplet continued to slowly decrease for approximately 15 minutes after this initial size loss, indicating an additional component leaving the droplet.

**Figure S8.**
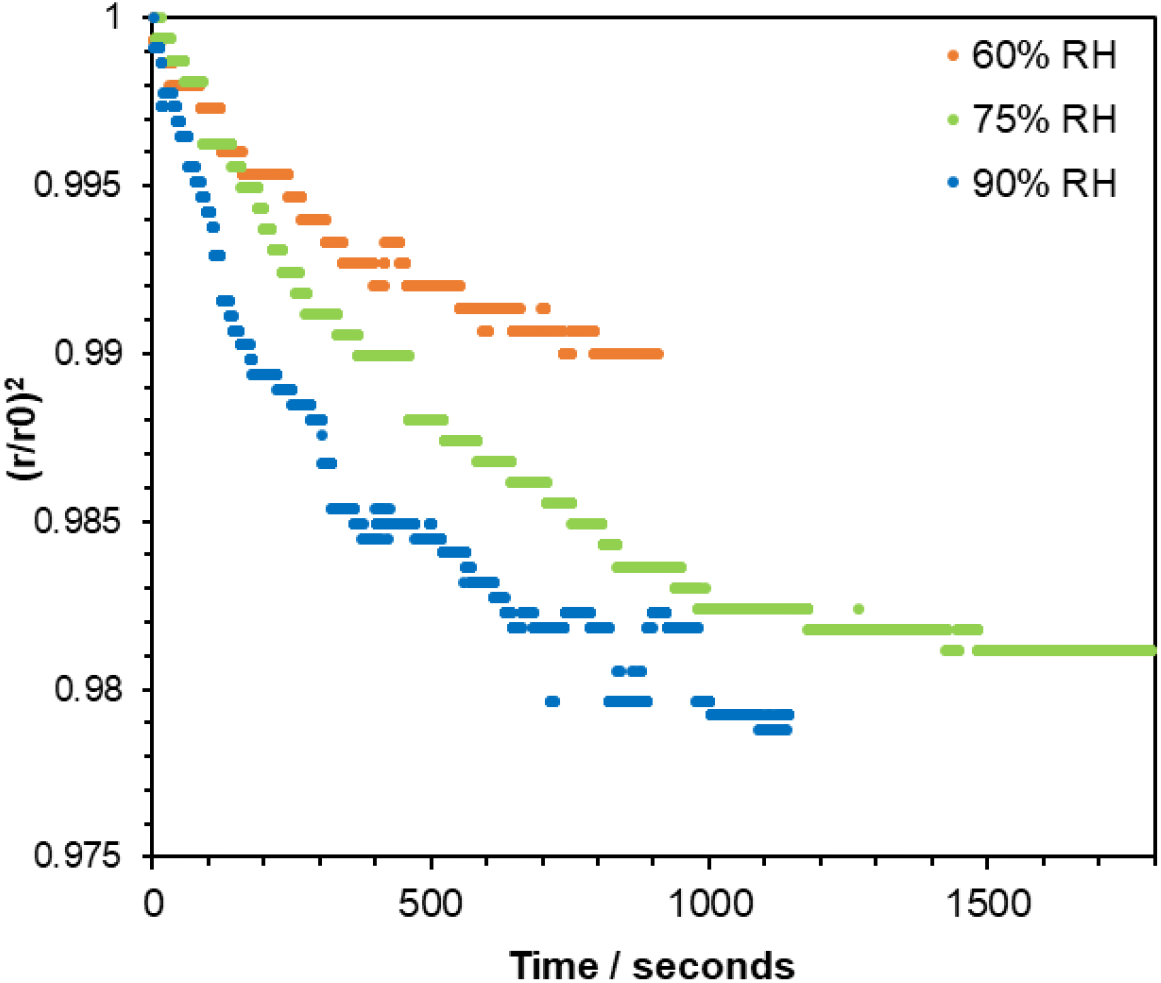
CO2 loss from sodium bicarbonate droplets. CO_2_ loss driven size change in airborne droplets. The evaporation of droplets of a 2:1 mass ratio of NaCl-NaHCO3 (to a final MFS of 0.08) was measured in the CK-EDB at 60% RH (orange), 75% RH (green) and 90% RH (blue). Radius is normalised to a point after the water has evaporated, allowing comparison of the radius change caused by CO_2_ loss.

**Figure S9.**
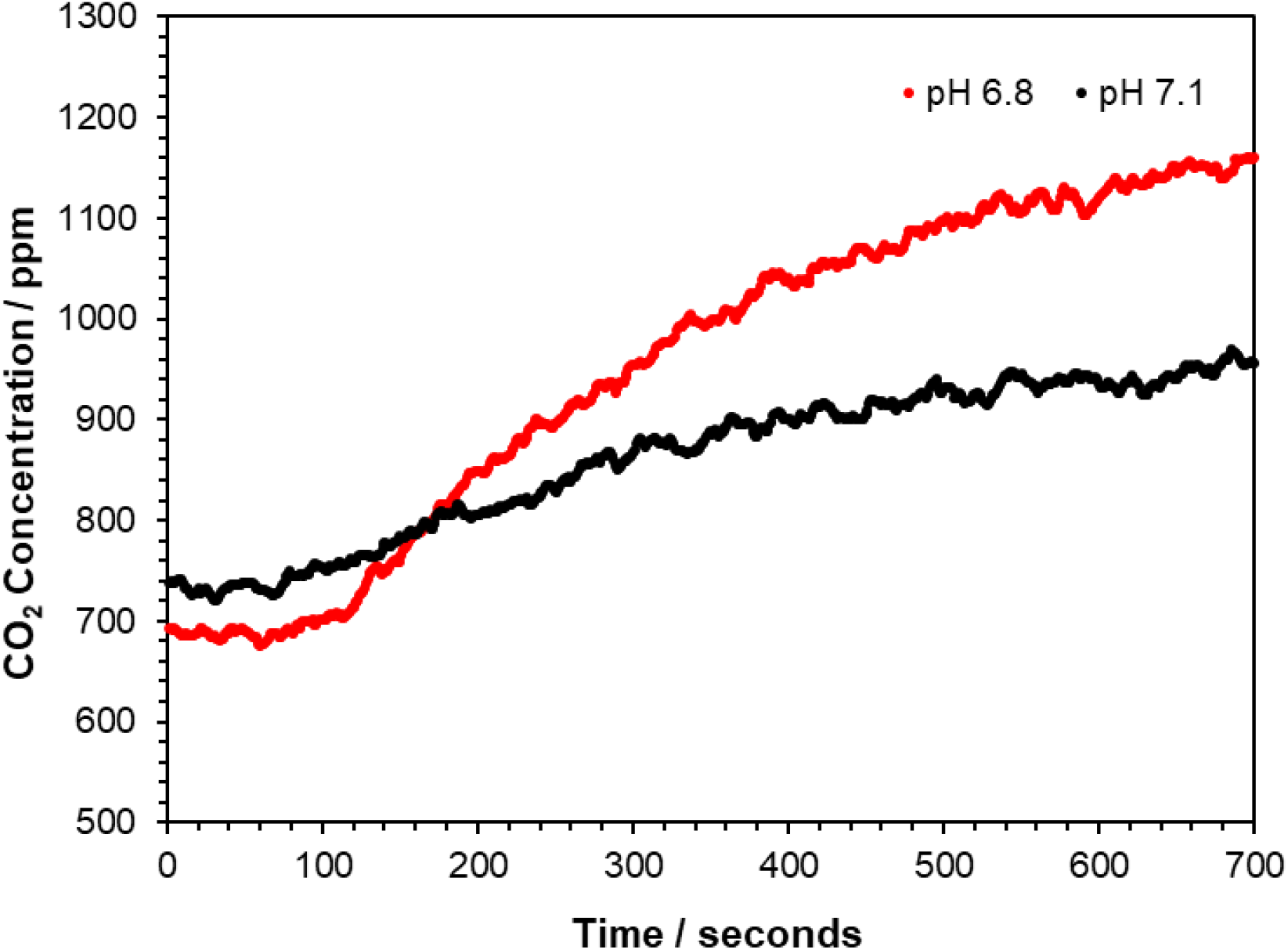
CO2_(g)_ Concentration Increase in an enclosed volume from MEM Nebulisation. Two solutions of MEM, one of pH 6.8 (red line) and one of pH 7.1 (black line) were nebulised into a 7-litre enclosed box, using a Collison nebuliser (50 PSI). Readings from a CO_2_ monitor placed within the box are reported here. The time 0 seconds marks the point at which the nebuliser was turned on. The overall increase in CO_2_ was found to be a function of the initial pH of the MEM.

**Figure S10.**
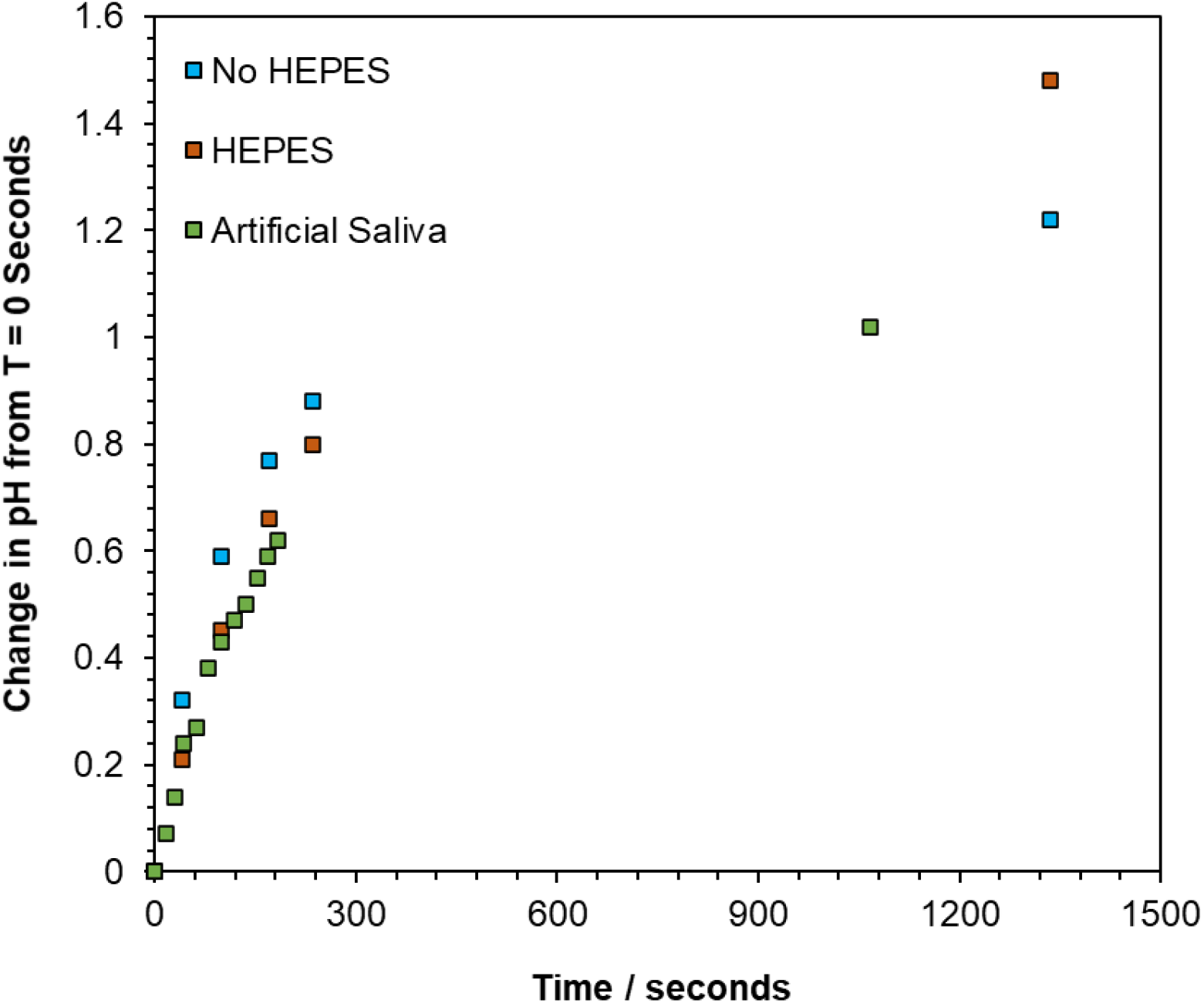
pH Increase in Nebulised Media. Three different solutions were placed into a nebuliser and the pH change in the nebuliser after turning the nebuliser on was measured. DMEM was nebulised both with (orange squares) and without (blue squares) HEPES. Artificial saliva was also nebulised (green squares). pH change is reported as the measured pH at the timepoint minus the pH of the solution before it was loaded into the nebuliser.

